# Longitudinal cross-species transmission of microbiomes and resistomes across farmers, animals and environment

**DOI:** 10.64898/2026.05.06.26352545

**Authors:** Jiabao Xing, Zhiying Xu, Yichao Zhang, Hang Zhang, Leyin Zheng, Meng Zhang, Wei Guo, Jianhua Liu, Yushan Pan, Junkai Zhang, Zhijun Jie, Guy Baele, Changchao Li, Alaric W. D’Souza, Jinxin Zhao, Jian Li, Tianmu Chen, Hua Wu

## Abstract

Understanding the acquisition and dissemination of microbiomes and antimicrobial resistance genes (ARGs) that circulate across human–animal–environment interfaces remains a central One Health challenge, largely because of complex ecological interactions and multiple confounding factors. Although occupational exposure is known to influence the microbiomes and resistomes of farmers, how environmental compartments involve in this system is unclear. Here, we conducted a one-year longitudinal study combining strain-resolved metagenomics (500 metagenomes) with isolate-based whole-genome sequencing (28 isolates) in an ecologically managed, antibiotic-free farming ecosystem spanning animals, farmers, environmental compartments and non-exposed individuals. Assembling 6,075 species-level genomes, we show that animal-associated occupancy reshapes the microbiome and resistome of occupationally exposed farmers and their surrounding environments. Animals and their associated habitats formed the dominant interface for both strain sharing and ARG dissemination across connected ecological compartments, whereas village residents and surrounding river samples - used as ecological controls - showed limited integration into this sharing network. Tracking a frequently shared lineage further revealed within-lineage genetic turnover together with selection-consistent changes following cross-species spread, suggestive of ecological selection across hosts and habitats. Finally, we identify *Klebsiella pneumoniae* as the most widespread ESKAPE pathogen in this ecosystem, with repeated occurrence across animal, human and environmental compartments, consistent with a neglected but clinically critical broad profile of ecological generalist. Together, these findings identify animals as central interfaces for microbiome and resistome sharing and show how agricultural ecosystems can sustain circulation of opportunistic pathogens and resistance determinants across human–animal–environment interfaces even in the absence of routine antibiotic use.

## Main

Occupational and environmental exposures in farming systems are increasingly recognized as major forces shaping the composition and functional capacity of the human microbiome and resistome, particularly in farming systems where humans, animals and their shared surroundings are in frequent contact^1–9^. In conventional livestock production, widespread antibiotic use has turned agricultural settings into important reservoirs and amplification hubs for antimicrobial resistance (AMR), while close and repeated contact between humans, animals and their shared surroundings can facilitate microbial exchange across ecological boundaries^1,3–5,10^. Yet microbial disruption in farming systems is not restricted to antibiotic-intensive settings. Transitions from non-agricultural to agricultural environments can rapidly alter both gut- and skin-associated microbial communities, together with their linked resistomes^2,5,11^. Once established, these shifts may persist in human hosts and in surrounding habitats, creating ecological reservoirs that sustain long-term transmission^12^.

Despite growing recognition of these farming-associated ecological linkages, several fundamental questions remain unresolved^13^. In antibiotic-free, free-range systems, it is still unclear whether microbiome and resistome sharing occurs to a measurable extent, how persistent such exchange is over time, and whether it is driven predominantly by direct human–animal contact or by indirect transmission through soil, water and farm-associated surfaces. Previous studies have reported microbial and resistome overlap between humans and animals, as well as occupation-associated signatures in farmers^4,5,8^ , but these insights are derived largely from cross-sectional or sparsely sampled datasets. As a result, they provide only static snapshots of a fundamentally dynamic process, limiting inference about transmission timing, persistence and directionality^14^. It therefore remains unresolved whether cross-host microbial movement simply reflects transient exposure, or instead represents sustained ecological exchange in which bacterial populations establish, adapt and diversify across host-associated and environmental niches^15^. These knowledge gaps are striking since nearly half a billion people globally are engaged in animal agriculture with chronic exposure to farm-associated microbial communities^16, 17^.

Resolving these dynamics is important not only for microbial ecology, but also for public health. Opportunistic pathogens are traditionally viewed through a clinical lens, yet many are increasingly detected in animals and surrounding environments, suggesting that non-human ecosystems may serve as reservoirs or intermediate niches for their maintenance and spread^18–20^. In particular, asymptomatic colonization is now recognized as a key mechanism enabling these pathogens to persist silently and subsequently seed difficult-to-control outbreaks^19^. Despite growing recognition that their ecology extends beyond hospitals, the non-clinical transmission dynamics of ESKAPE (*Enterococcus faecium, Staphylococcus aureus, Klebsiella pneumoniae, Acinetobacter baumannii, Pseudomonas aeruginosa and Entobacter* species) pathogens remain poorly resolved, particularly across asymptomatic human-animal-environment interfaces in agricultural systems.

Here, we established the OH-MicroDynamics (One-Health Microbiome Dynamics) project, a densely sampled longitudinal study conducted in an ecologically managed rural farming community without routine antibiotic use. Our sampling was preceded by six months of on-site observation and field characterization. By systematically sampling farmers, livestock, environmental reservoirs and non-farming residents living within the same ecosystem, we generated an integrated, high-resolution dataset encompassing microbiome, metabolome and resistome profiles together with strain-resolved genomic tracking (**Extended Fig. 1**). This framework enabled us to reconstruct the longitudinal transmission dynamics of microbial communities, resistance determinants, and ESKAPE pathogens across the farmer–animal–environment interface (**Extended Fig. 2**). We show that microbiome and resistome exchange is persistent and dynamic across hosts and habitats. Animals play a central role in exchange across environments and farmers, and we identify cross-species transmission events followed by signatures of adaptive divergence. We also uncover an unexpected ecological niche supporting cross-host colonization by *K. pneumoniae*. These findings provide a quantitively longitudinal One Health view on how microbial communities, resistance determinants and opportunistic pathogens circulate, establish and persist within a multi-interface farming ecosystem.

## Results

### The metagenomic study of OH-MicroDynamics

Our study was conducted in a remote rural village in Kaifeng, Henan Province, China, comprising fewer than 200 households, substantially below the provincial rural average (∼600 households) (**Extended Fig. 1**). Following extensive field reconnaissance and long-term ethnographic assessment, we identified an isolated ecological farm (hereafter referred to as the “ecosystem”) as the focal sampling site to investigate sustained ecological interactions among farmer, animal, and environmental microbiomes in the absence of routine antibiotic use. The site was selected for its minimal connectivity to surrounding agricultural and urban systems, representing a relatively self-contained microbial ecosystem with limited external anthropogenic perturbation^21^ (**Extended Fig. 1B**).

A detailed study site description is provided in the **Supplementary Note 1**. Within this ecosystem, we longitudinally tracked the nasal and gut microbiomes of all four resident farm workers. Animal sampling comprised gut microbiome profiles from each resident species: peacocks (*Pavo cristatus*), ostriches (*Struthio camelus*) and geese (*Anser cygnoides domesticus*), all sampled as adults to minimize age-related microbiome instability (see **Methods**; **Table S1**). Environmental samples were collected from key surrounding habitats, including the goose paddling pool, ostrich-associated soil, goose-associated soil, and a nearby river just beyond the ecosystem boundary (**Fig. 1; Extended Fig. 1**). As a paired community control, we concurrently collected nasal and fecal samples from demographically matched non-farming residents living in the same village but geographically separated from the farm ecosystem (**Extended Fig. 1**). All farmer and animal subjects were individually code-matched and followed longitudinally for one year, yielding 500 high-quality biological samples for deep shotgun metagenomic sequencing. We generated an average of 65.43 million microbial reads per sample.

**Figure 1.**
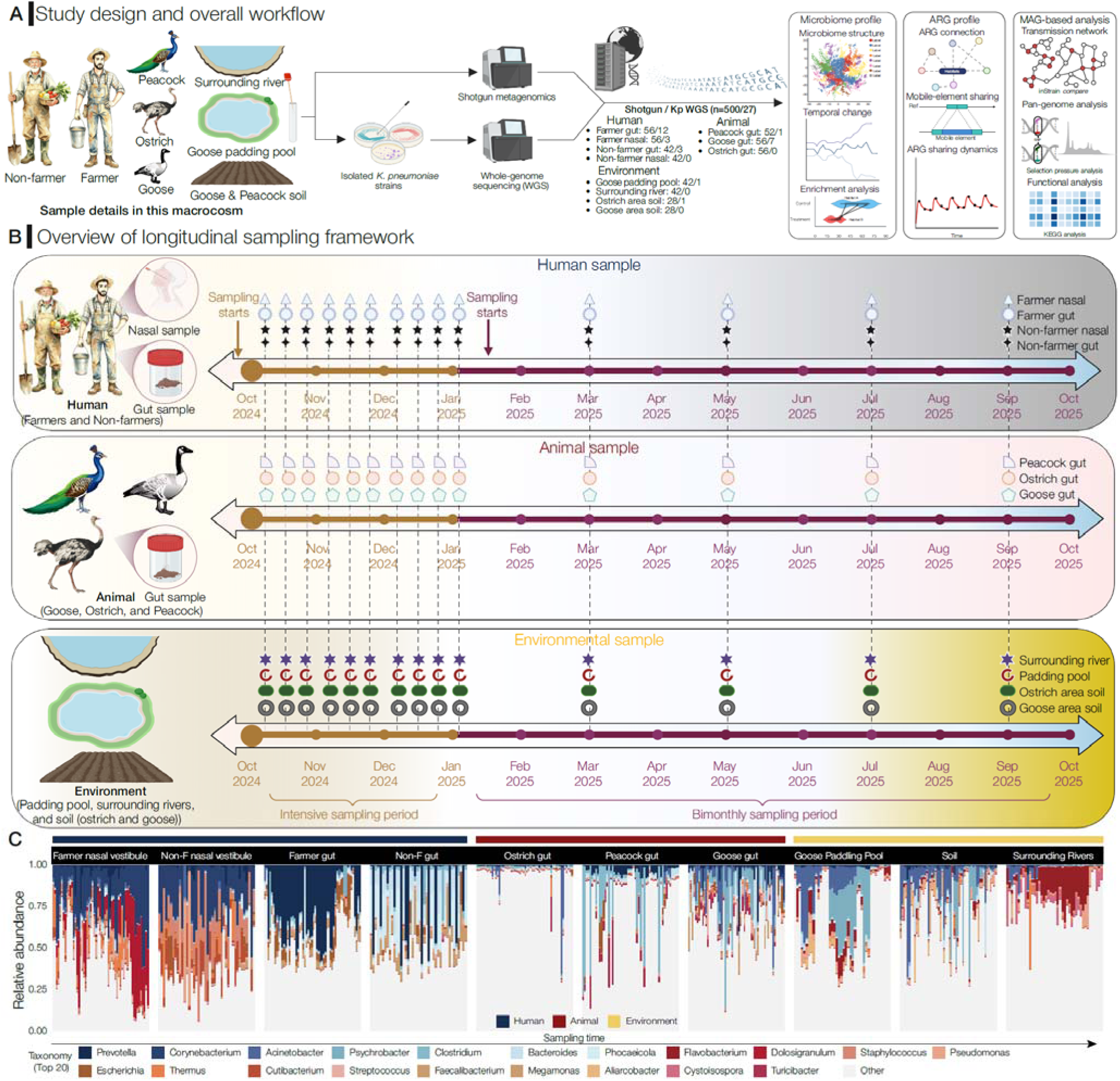
Study design, longitudinal sampling, and microbial composition across habitats. **A.** Schematic overview of the study design, including sampling objects, longitudinal sampling strategy, sequencing workflow, and analytical framework. All samples underwent metagenomic shotgun sequencing, while K. pneumoniae isolates recovered from corresponding samples were further subjected to whole-genome sequencing (WGS). The sample types and corresponding sample numbers are annotated in the figure. **B.** Schematic representation of the sampling scheme used for both initial and longitudinal shotgun metagenomic sequencing. Three modules correspond to distinct habitats within the ecosystem, i.e., farmer, animal, and environmental samples. Each icon along the timeline represents one sampling event (note that the icons do not indicate sample numbers). The full sampling campaign spanned one year and was divided into two phases: intensive sampling period (October 2024-January 2025) and bimonthly sampling period (February-October 2025). **C.** Relative abundance of the top 20 bacterial families across habitats. Each facet represents a specific habitat, with bars arranged chronologically to illustrate temporal variation.

### Animal habitation drives microbiome convergence across farmers and their environment

We first assessed sequencing depth adequacy by estimating γ-diversity^22^ across habitats. Sampling completeness reached a median of 93.1% (IQR 89.5-95.0%), indicating that most of the detectable diversity was captured (**Fig. 2B**). Microbial diversity varied significantly among hosts and environments (**Fig. 2A; Supplementary Fig. S1**). Farmer nasal samples exhibited lower α-diversity (indexed by richness and Shannon) than gut samples (*P* < 0.0001), whereas both nasal and gut microbiomes of farmers showed higher α-diversity than those of non-farmers (*P* < 0.05). Among animal hosts, ostrich gut microbiomes displayed the highest α-diversity, exceeding that of geese and peacock (*P* < 0.001), a pattern mirrored in their associated habitat soils (*P* < 0.001). Bray–Curtis dissimilarities revealed strong compositional stratification across hosts and environments, with human microbiomes clearly separated from those of animals (dissimilarity = 0.7-0.9) (**Fig. 2C; Extended Fig. 3B**). In contrast, goose gut, goose-associated soil, and paddling pool microbiomes were more similar (dissimilarity = 0.37–0.56). Within humans, nasal–gut dissimilarity (0.47–0.61) remained lower than inter-species distances. Environmental samples occupied an intermediate niche, bridging animal and human microbiomes. Notably, farmers showed reduced dissimilarity to animal-associated environments (0.78–0.85) relative to non-farmers, indicating that long-term occupational exposure promotes cross-host microbiome convergence.

**Figure 2.**
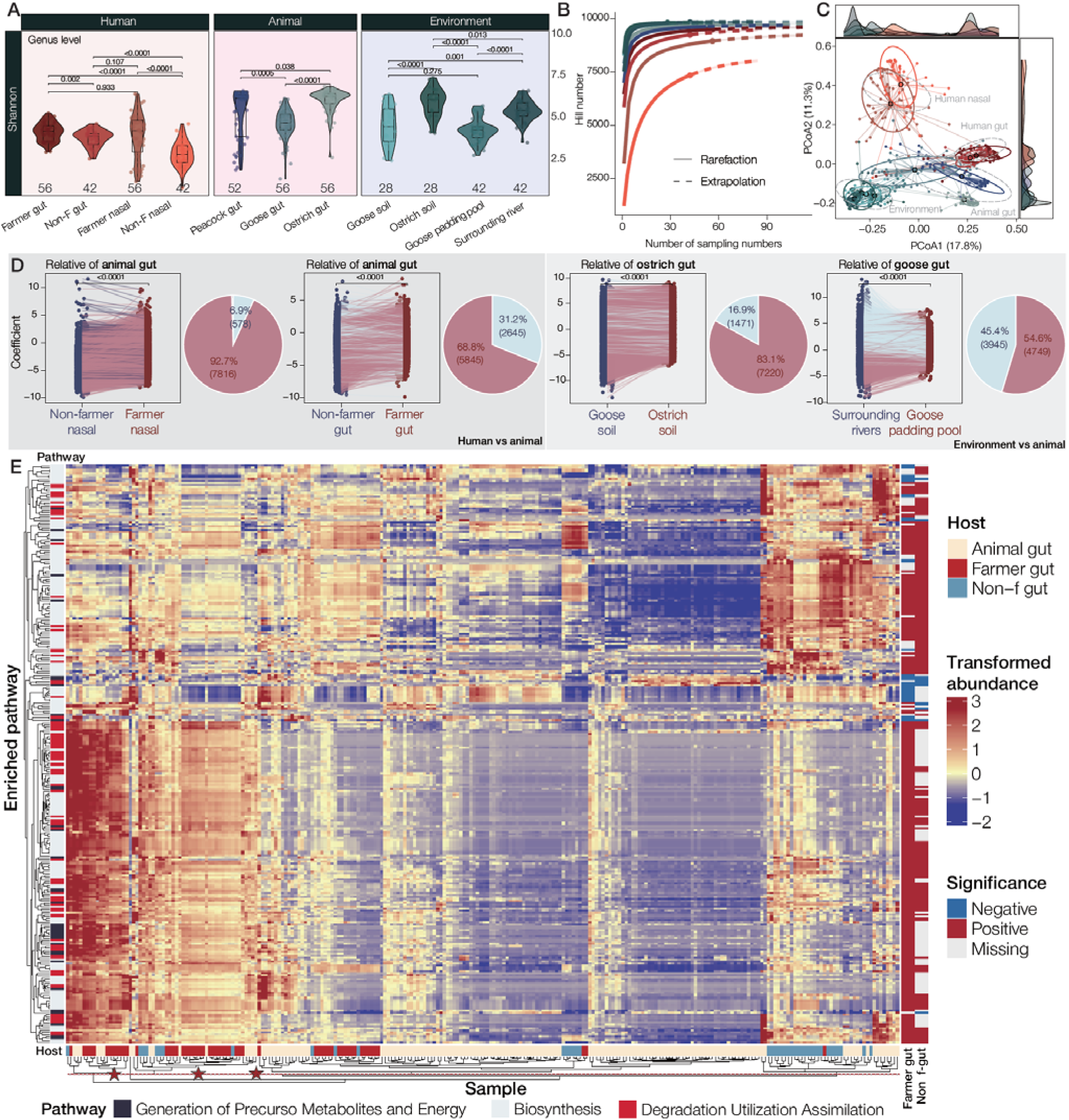
Differentiation and interconnection of metabolic potential between human, animal, and environmental microbiomes. **A.** Violin plots showing genus-level α-diversity (Shannon index) across habitats. Samples are grouped by major habitat categories (human, animals, and environment) and further stratified by specific sampling sites. Horizontal lines indicate median values. P values were calculated using the two-tailed Wilcoxon rank-sum test, with subsequent Benjamini–Hochberg correction for multiple hypotheses. **B.** Sample-size-based rarefaction (solid lines) and extrapolation (dashed lines) curves of genus-level richness (q = 0) for each habitat, with shaded areas indicating 95% confidence intervals. Curves illustrate sampling completeness and projected diversity beyond the observed samples. **C.** PCoA based on Bray-Curtis dissimilarity of genus-level microbial community composition across habitats. Shaded ellipses represent 95% confidence intervals, and marginal density plots illustrate sample distributions along the first two PCoA axes. **D.** Comparison of differential abundance coefficients across habitat pairs with statistical significance inferred using MaAsLin2. Left: paired scatterplots showing regression coefficients for individual bacterial taxa across comparisons. Lines connect paired coefficients for the same taxa. Right: pie charts summarize the proportion of significantly enriched bacterial features in each habitat. Statistical significance was determined using Benjamini-Hochberg-adjusted P values. **E.** Heatmap showing microbial metabolic pathways that differ significantly in relative abundance across host groups, identified using MaAsLin2 (adjusted P < 0.05). Rows represent metabolic pathways and columns represent samples. Color intensity indicates transformed relative abundance from low (blue) to high (red). The left-hand annotation bar indicates pathway functional categories (generation of precursor metabolites and energy, biosynthesis, and degradation/utilization/assimilation). The right-hand annotation bar indicates the direction of significant differential enrichment. Red stars highlight pathways showing evidence of host mixing across groups.

To further evaluate this pattern, we performed a statistical enrichment analysis of host–environment overlap using MaAsLin2^23^. Farmer nasal and gut microbiomes exhibited stronger enrichment toward animal-associated communities than non-farmer microbiomes, whereas animal-living environments (soil and water) had greater compositional similarity to their host’s gut microbiome (*P* < 0.001; **Fig. 2D**). Repeated-measures PERMANOVA further identified sample type, occupational exposure in humans, and proximity to animal habitats in environmental samples as major correlates explaining microbiome variation (**Extended Fig. 3A**). Together, these findings suggest that sustained animal habitation may reshape nearby environmental and human microbiomes, generating a shared ecological continuum that partially transcends host and habitat boundaries. Importantly, this convergence extended beyond taxonomic composition. HUMAnN3-based functional potential profiling further showed that farmer gut and nasal microbiomes were more similar to animal-associated communities than those of non-farmers, with cross-habitat overlap particularly evident in pathways related to degradation, utilization, assimilation, and biosynthesis^24^ (**Supplementary Note 3**; **Fig. 2E; Extended Fig. 3C; Supplementary Fig. S2**).

Thus, long-term farming occupational exposure appears to promote parallel convergence in both microbiome composition and metabolic potential^3,25^.

### Resistome composition is hierarchically structured across the farming ecosystem

To characterize overall resistome composition across the ecosystem, we profiled antibiotic resistance genes (ARGs) in all shotgun metagenomes using ARGs-OAP v3.0^26^. Overall, ARG abundance and diversity followed a clear habitat hierarchy, with human gut samples showing the highest levels, followed by animal guts and then environmental compartments (**Extended Fig. 4A, D**). Farmers harbored more abundant and diverse resistomes (reflected by richness, Shannon diversity, and average variability degree; AVD - see **Methods**) than non-farmers at both gut and nasal sites (*P* < 0.001). Among animal hosts, ostrich guts carried significantly higher ARG Shannon diversity than goose guts, a contrast that was also reflected in their corresponding habitat soils (*P* < 0.001; **Extended Fig. 4A**). This pattern suggests that soil resistomes partially mirror host-associated differences in the resident animal communities. Bipartite network analysis further supported this interpretation (*P* < 0.001; **Fig. 3B**). Strong positive associations linked animal guts with their immediate environments. Farmer gut resistome emerged as a major terminal node. By contrast, human nasals - particularly those from non-farmers - displayed multiple negative associations **(Fig. 3B)**. Consistent patterns were observed in the Procrustes shape analysis, which quantified the coupling between microbiome and resistome structures (**Fig. 3C; Extended Fig. 4E**). Although the two assemblages were significantly concordant overall (m^2^= 0.3791, P < 0.001, PROTEST), the strength of coupling varied by habitat. Animal guts and goose-associated soils showed the lowest residuals, indicating tight structural correspondence, whereas ostrich soils appeared comparatively less stable. In contrast, nasal samples, especially those from non-farmers, showed high residuals, suggesting weaker microbiome–resistome coupling^27^.

**Figure 3.**
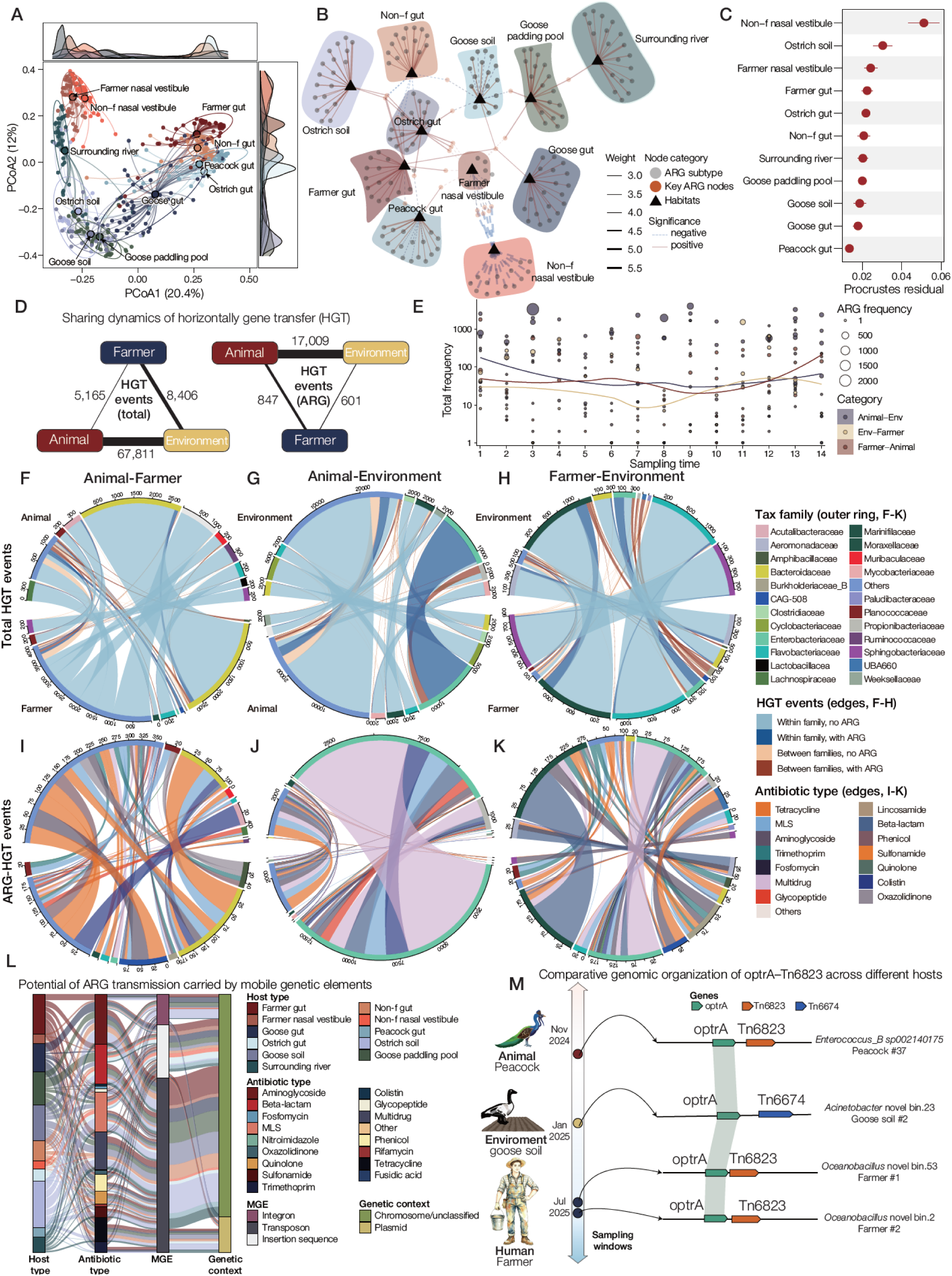
Spread and horizontal gene transfer of ARGs across human, animal and environmental microbiomes. **A.** Principal coordinate analysis (PCoA) based on Bray-Curtis dissimilarity of resistome composition across habitats. Each point represents a sample and colors denote sampling habitats. Shaded ellipses indicate 95% confidence intervals, and marginal density plots illustrate sample distributions along the first two axes. **B.** Bipartite network linking ARG subtypes and habitats. Triangles represent habitat nodes and circular nodes represent ARG subtypes. Edge width reflects the strength of association between ARGs and habitats, whereas edge style indicates statistical significance. **C.** Procrustes residuals quantifying the degree of concordance between microbiome composition and resistome profiles across habitats. Lower residual values indicate stronger microbiome-resistome coupling (see **Extended** Fig. 4E for Procrustes analysis). **D.** Total HGT events detected across habitats (left) and HGT events involving ARGs (right). **E.** Temporal dynamics of HGT event frequencies across sampling time points. Each point represents a sample and lines indicate smoothed trends for different host-environment pairings. **F-H.** Chord diagrams illustrating all detected HGT events between host groups: animal-farmer (F), animal-environment (G), and farmer-environment (H). The outer rings represent MAG family assignments, with segment size proportional to the number of fragment-sharing events involving each family. Edges represent shared DNA fragments identified across distant MAGs and are colored by family pairs and ARG-encoding status. **I-K.** Chord diagrams showing ARG-encoding HGT events between host groups: animal-farmer (I), animal-environment (J), and farmer-environment (K). Edges are colored according to the antibiotic class of the encoded ARG. For fragments encoding ARGs from multiple antibiotic classes, each class is represented by a separate edge. **L.** Sankey diagram illustrating the co-occurrence of ARGs and mobile genetic elements (MGEs) within 10 kb genomic windows on the same contig. Flows represent connections between habitat type, antibiotic class, MGE type, and genetic context. **M.** Comparative genomic organization of the optrA-Tn6823 resistance cassette identified across animal, environmental, and human-associated microbial genomes. Conserved genetic structures carrying optrA were detected in peacock gut, goose soil and human-associated genomes across multiple sampling periods, indicating potential cross-host dissemination mediated by MGEs.

We next examined ARG composition in detail. Since no routine antibiotics were used in this ecosystem in documentation spanning two years, this system provided a reasonable natural experiment for assessing ARG composition and temporal fluctuation without direct antibiotic use (see **Methods**). Farmer samples harbored the largest number of high-risk ARGs (Rank I gene defined in ref ^28^), followed by goose, peacock and ostrich samples (**Extended Fig. 4F and Extended Fig. 5A–C**). Notably, the carbapenemase gene *bla*_NDM_ was frequently detected in farmer and soil samples (**Extended Fig. 5C**). In addition, several antibiotic classes - including vancomycin, chloramphenicol, novobiocin, streptothricin, rifamycin and florfenicol resistance determinants - were consistently enriched in animal guts (**Extended Fig. 4C**). Of these, only florfenicol is licensed for veterinary use, whereas the remaining classes are primarily reserved for human clinical settings. To investigate temporal resistome dynamics, we tracked the 10 most prevalent ARG subtypes across sampling timepoints (**Extended Fig. 5B, D**). Despite being widely distributed across the ecosystem, human-associated ARGs showed pronounced temporal variability, whereas animal gut and environmental resistomes remained comparatively stable over time.

### Horizontal ARG dissemination is concentrated along the animal-environment interface

To directly quantify ARG dissemination across the ecosystem, we extended an established framework for identifying *horizontal gene transfer (HGT)* events and tracked the temporal dynamics of ARG-associated transmission^29^. We first benchmarked several widely used metagenomic binning strategies and found the multi-sample binning approach outperformed alternatives for our dataset (**Supplementary Note 2; Extended Fig. 6**)^29^. Using this strategy, we reconstructed a total of 29,838 medium- and high-quality metagenome-assembled genomes (MAGs), which subsequently yielded 6,075 species-level genome bins (SGBs; **Table S2**).

Using pairwise comparisons, we identified 81,382 total HGT events, of which 18,457 involved ARG-associated transfer (**Fig. 3D**). Among sharing events, direct HGT between animals and the environment overwhelmingly predominated (N = 67,811, 83.33%), followed by farmer–environment pairs (N = 8,406, 10.33%) and animal–farmer pairs (N = 5,165, 6.35%). Restricting the analysis to ARG-associated transfer revealed a similar but even more polarized pattern: ARG sharing was dominated by animal–environment pairs (N = 17,009), whereas direct ARG transfer between farmers and the environment (N = 601) and between farmers and animals (N = 847) occurred at much lower frequencies. Although the absolute number of horizontally transferred fragments fluctuated over time, the overall ranking of transmission routes remained stable across sampling timepoints (**Fig. 3E**).

We next compared total HGT and ARG-associated HGT events involving farmers or non-farmers relative to other compartments (**Extended Fig. 7C**). Transmission between farmers and animals occurred at approximately twice the frequency observed for non-farmers, and the proportional enrichment was even greater for ARG-associated events. This contrast was even more pronounced in farmer–environment comparisons. These results are consistent with the hypothesis that sustained occupational exposure increases opportunities for horizontal exchange and elevates the frequency of ARG sharing. Though most HGT events occurred within the same bacterial family, transmission patterns remained strongly pair-specific. For example, farmer–animal transfer events were dominated by members of the *Bacteroidaceae* and *Marinifilaceae*, whereas farmer–environment transfers showed a similar overall trend but primarily involved *Flavobacteriaceae*, *Moraxellaceae* and *Sphingobacteriaceae* (**Fig. 3F-H**). By contrast, animal–environment transmission was dominated by *Cyclobacteriaceae*, *Enterobacteriaceae* and a broad set of additional taxa. This compartment specificity was also reflected in ARG-associated transfer profiles. ARGs exchanged between farmers and animals were primarily associated with tetracycline and colistin resistance (e.g., *tet(X)* and *mcr-1*), whereas farmer–environment transmission was enriched in beta-lactam and multidrug resistance determinants. In contrast, multidrug resistance genes overwhelmingly dominated ARG transfer between animals and the environment, followed by beta-lactam, tetracycline and colistin resistance genes (**Fig. 3I-K**).

To illustrate the spatiotemporal connectivity and ecological breadth of ARG dissemination, we identified a representative case involving the oxazolidinone resistance gene *optrA* (**Fig. 3M**). In November 2024, we first detected a Tn6823-associated *optrA* gene in an *Enterococcus_B* sp002140175 MAG. In January 2025, the same resistance determinant was subsequently detected in a goose-soil *Acinetobacter* novel bin.23, although in this case it was associated with Tn6674. Thereafter, we detected Tn6823-associated *optrA* in gut samples from two workers at successive timepoints, in both cases carried by *Oceanobacillus*-affiliated strains. These three hosts were taxonomically distinct and were detected at different timepoints with 99.5% average nucleotide identity (ANI), providing direct evidence that ARGs can disseminate repeatedly across the ecosystem even in the absence of recorded antibiotic administration. This observation motivated us to further characterize ARG mobility (**Extended Fig. 7A-B**). Two contrasting distributional patterns emerged. First, insertion sequences were the most prevalent MGEs across all 11 sample categories, whereas transposons were less frequent and integrons were consistently the least common, in which rank order was highly conserved across habitats. Second, among loci where ARGs co-occurred with MGEs, transposons were the dominant carriers. This asymmetry suggests that the most prevalent MGEs in the ecosystem are not necessarily those most directly involved in ARG mobilization. In addition, the frequency of ARG–MGE co-occurrence varied by antibiotic resistance category, with MLS, aminoglycoside, tetracycline and beta-lactam resistance genes the predominant classes co-occurring with MGEs across farmer, animal and environmental compartments (**Fig. 3L**).

These findings show that HGT events and ARG dissemination preferentially concentrated along defined ecological interfaces. Environment is a major exchange interface, with animal–environment interactions accounting for most total HGT and ARG-associated transfer, while occupational exposure increases farmers’ connectivity to the sharing network compared with non-farmers (**Fig. 3M**).

### Strain sharing centers at animal-environmental interfaces and secondarily extends to exposed farmers

Taxonomic profiling with MetaPhlAn v4 suggested extensive strain-level novelty (**Fig. 1C**). This pattern was consistent with our genome-resolved analysis, in which 3,904 of 6,075 SGBs represented potentially novel species-level diversity (64.26%), underscoring the substantial unexplored microbial diversity captured across this farming ecosystem (**Fig. 4B; Supplementary Note 4**). Notably, microbiome novelty was unevenly distributed across exposure groups and body sites. In nasal samples, farmer-associated microbiomes contained a markedly higher fraction of novel MAGs than non-farmer samples (Δ = 0.217, *P* = 1 × 10^-4^), and this difference remained robust across abundance thresholds. In gut samples, farmers also showed a modest but consistent increase in the fraction of novel MAGs compared with non-farmers (Δ ≈ 0.028), although this trend did not reach statistical significance across filtering thresholds (P ≈ 0.06; **Extended Data Fig. S8G–N**).

**Figure 4.**
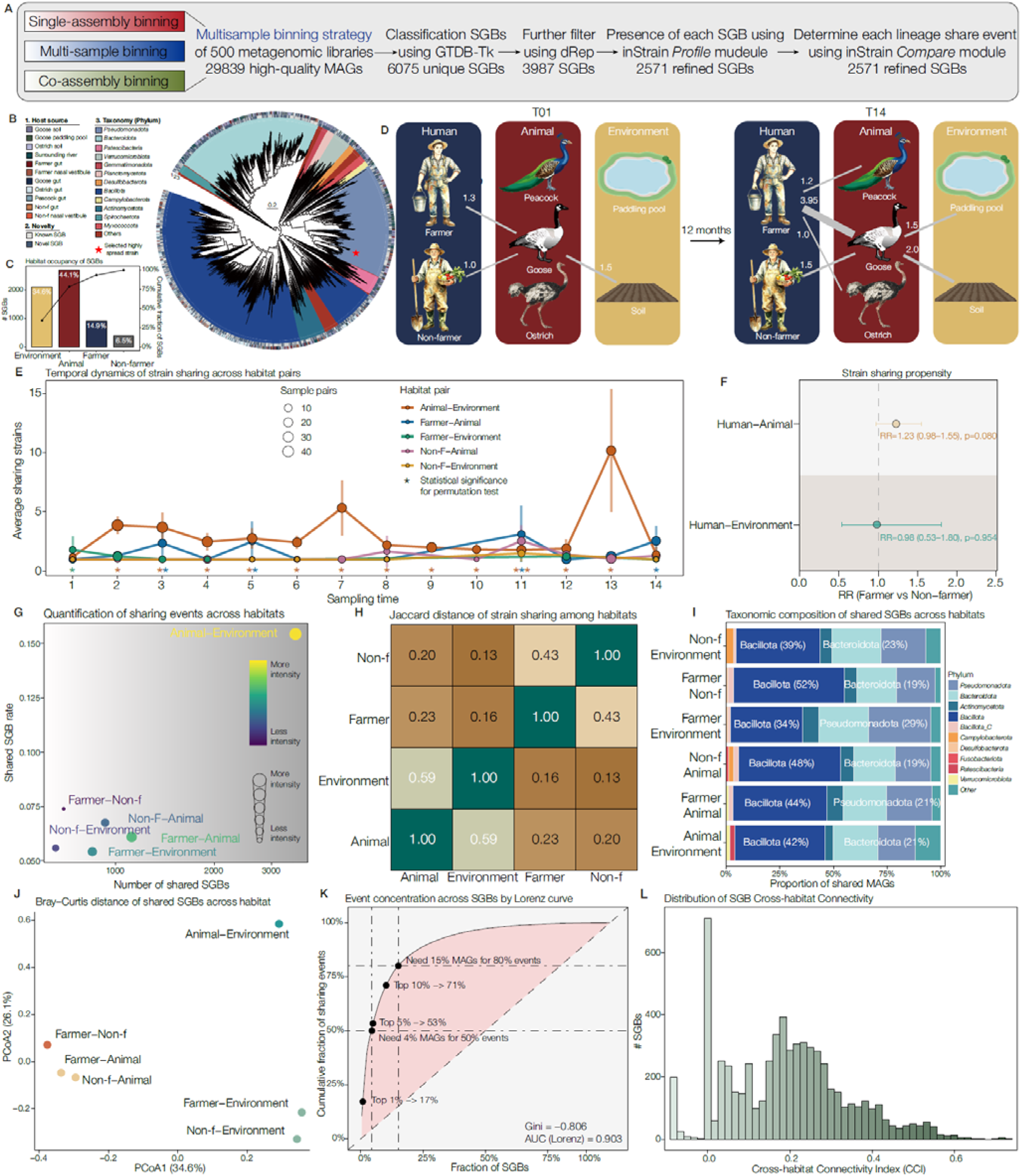
Longitudinal transmission dynamics across habitats (hosts) during sampling periods. **A.** Analytical workflow for identifying strain-sharing events across metagenomic samples. Three binning strategies were initially considered, and the final analysis was based on a multi-sample binning strategy applied to 500 metagenomic libraries^31^. **B.** Phylogenetic overview of recovered SGBs. Phylogeny shows the taxonomic placement of SGBs, with outer annotation tracks indicating habitat origin, phylum-level taxonomy, and genomic novelty. **C.** Habitat occupancy of recovered SGBs across the four major ecological groups (environment, animal, farmer, and non-farmer). Bars indicate the number and proportion of SGBs detected in each group, and the line shows the proportion of all recovered genomes represented in each habitat category. **D.** Conceptual summary of cross-habitat strain sharing events at the beginning (T01) and end (T14) of longitudinal follow-up. Nodes represent the four major ecological groups and their constituent habitats, and edge widths schematically illustrate the relative intensity of lineage sharing between groups over time. **E.** Temporal dynamics of strain sharing across habitat interfaces. Lines show the average number of shared strains per sample pair across 14 sampling time points. Point size indicates the number of contributing sample pairs. Insets summarize the cumulative contribution of farmers versus non-farmers to human-animal and human-environment strain sharing. Asterisks indicate significance from permutation-based tests comparing observed lineage-sharing intensity against opportunity-adjusted null expectations. **F.** Relative propensity for strain sharing involving farmers versus non-farmers. Points represent risk ratios (RRs) estimated from a negative binomial regression model of shared-strain counts with human type, habitat interface, and their interaction. Error bars indicate 95% confidence intervals. RR > 1 indicates greater strain-sharing propensity for farmers relative to non-farmers within the corresponding interface. **G.** Quantification of strain-sharing intensity across major habitat pairs. Each point represents one habitat-pair comparison, plotted by the total number of shared SGBs (x axis) and the number of shared SGBs normalized by the number of possible inter-group sample pairs (y axis). Point size and color both scale with sharing intensity. **H** Pairwise Jaccard similarity of SGB occurrence across the four major ecological groups, calculated from stringent SGB presence profiles. Values indicate the proportion of shared SGBs relative to the union of SGBs detected in each habitat pair. **I.** Phylum-level composition of SGBs shared across habitat pairs. Stacked bars show the relative contribution of dominant bacterial phyla to the pool of shared SGBs for each pairwise habitat comparison. **J.** PCoA of habitat-pair-specific shared-SGB composition based on Bray-Curtis dissimilarity. Each point represents one habitat-pair comparison. **K.** Lorenz curve showing the concentration of strain-sharing events across SGBs. SGBs were ranked by their contribution to weighted sharing events, and the cumulative fraction of sharing events is plotted against the cumulative fraction of SGBs. This analysis highlights the disproportionate contribution of a small subset of SGBs to overall strain sharing. **L.** Distribution of the cross-habitat connectivity index (CCI) across SGBs. CCI integrates four components: breadth of occurrence across sampling sources, span across the four major ecological groups, prevalence in human-associated samples, and contribution to cross-group sharing events. Higher CCI values indicate SGBs with greater potential to connect multiple habitats and host types (see **Methods**).

To characterize how microbial lineages disseminate across hosts and habitats in this farm ecosystem, we quantified strain sharing on 2,571 high-confidence SGBs identified using an inStrain-based framework (**Fig. 4A**). We found that strain sharing was widespread across the ecosystem, but was also primarily concentrated at the animal–environment interface (**Fig. 4C, G**). Intriguingly, this pattern of strain sharing strengthened over time (**Fig. 4D**). At the beginning of sampling (T01), only limited sharing was detected, most notably between geese and farmers or goose soil. By the final sampling (T14), the number of shared strains involving geese had increased substantially. Farmers and geese shared an average of 3.95 strains, farmer–peacock sharing reached 1.2, and geese also shared 1.5 and 2.0 strains with the paddling pool and goose soil, respectively (**Fig. 4D**). Notably, direct human–environment sharing remained absent at both early and late snapshots. Although the residence time of different animal species in the ecosystem was not identical, geese consistently showed a greater capacity to share strains with humans than the other animal hosts, pointing to species-specific differences in their ability to seed or acquire microbiota across host boundaries. This early prominence of geese, however, did not fully capture the long-term dynamics observed across the full sampling period. When all 14 timepoints were considered together, animal–environment sharing remained consistently higher than any other habitat pair from T01 to T13, followed by farmer–animal sharing. Non-farmer controls exhibited minimal strain sharing with farm-associated compartments (**Fig. 4E-F**). Using negative binomial regression, we found that farmers showed a higher relative risk for strain sharing with animals compared to non-farmers, whereas differences at the human–environment interface were modest (**Fig. 4F**). We tested whether the elevated sharing observed at the animal–environment interface could be explained by unequal sampling opportunities. Permutation analyses under both global and time-stratified null models showed that the enrichment of strain sharing remained significant across both soil- and water-associated habitats, indicating that the observed pattern was not driven by pairwise sampling structure or study design (**Extended Fig. 9C-D**). Together, these results support the conclusion that strain sharing is preferentially concentrated in animal-associated habitats, consistent with repeated accumulation and redistribution of shared lineages in environments directly occupied by animals.

We then assessed whether shared lineages were compositionally similar across habitat interfaces. Jaccard overlap and Bray–Curtis dissimilarity showed that animal and environmental compartments were the most similar in shared-strain composition, whereas human-associated interfaces remained more distinct (**Fig. 4H–J**). Shared SGBs across habitat pairs were dominated by *Bacillota* and *Bacteroidota*, although their relative contributions varied among interfaces (**Fig. 4I**). Ordination analysis recapitulated the same gradient at the community level, with animal–environment shared pools clearly separated from farmer- and non-farmer-associated comparisons (**Fig. 4J**). Together, these results suggest that cross-habitat sharing is structured not only by contact opportunity, but also by the ecological capacity of specific lineages to persist across multiple compartments. We next asked whether strain sharing was broadly distributed across lineages or concentrated within a subset of highly connected taxa. Lorenz-curve and the cross-habitat connectivity index (CCI) analyses jointly showed a strongly skewed distribution, corroborating that a small subset of SGBs accounts for a disproportionate fraction of all strain-sharing events (**Fig. 4K-L**)^30^. Together, these results indicate that cross-habitat strain sharing is disproportionately driven by a limited subset of ecologically well-connected lineages.

Overall, these findings identify animal-associated environments as the primary exchange zone for strain sharing in this ecosystem. Although occupational exposure integrates farmers into this network, their level of involvement remains markedly lower than that observed at the animal–environment interface. This pattern is concordant with the sharing structure inferred from HGT events and ARG dissemination.

### Evolutionary signal to adapt to new hosts for an extensively disseminating strain

To investigate why only a subset of strain lineages achieved extensive dissemination across the farm ecosystem (**Fig. 4K-L**), we focused on the SGB with the highest number of inferred sharing events: *E. coli GF12EGFS_2_bin.26*. This lineage provided a tractable model for resolving how a successful disseminating strain moves across hosts and habitats, and whether such spread is accompanied by adaptive change in response to host- and habitat-specific selection.

As a representative example of combined capabilities of our study design and metagenomic pipeline to reconstruct complex transmission chains, we resolved the phylogenetic landscape of this SGB within the ecosystem and identified a terminal cluster with well-supported transmission links (**Fig. 5A-C**). The reconstructed transmission chain suggested that the strain was initially carried by farmer #3 at T01, subsequently transmitted to farmer #4 at T02, and by T04 had already undergone broader dissemination, including non-farmer #2 and the surrounding river. At T06, the strain was transiently detected in the nasal sample of non-farmer #1. The lineage persisted continuously in the guts of farmer #3 and farmer #4 and was later detected in farmer #1 and farmer #2 at T11. Although it was not detected at T12, it reappeared in farmer #3 and farmer #4 at T13 and T14, consistent with sustained persistence and repeated re-emergence rather than one-off introduction events (**Fig. 5A**). However, we note that intermittent non-detection may reflect limits of metagenomic detection sensitivity, and the lineage may have persisted at low abundance below the detection threshold.

**Figure 5.**
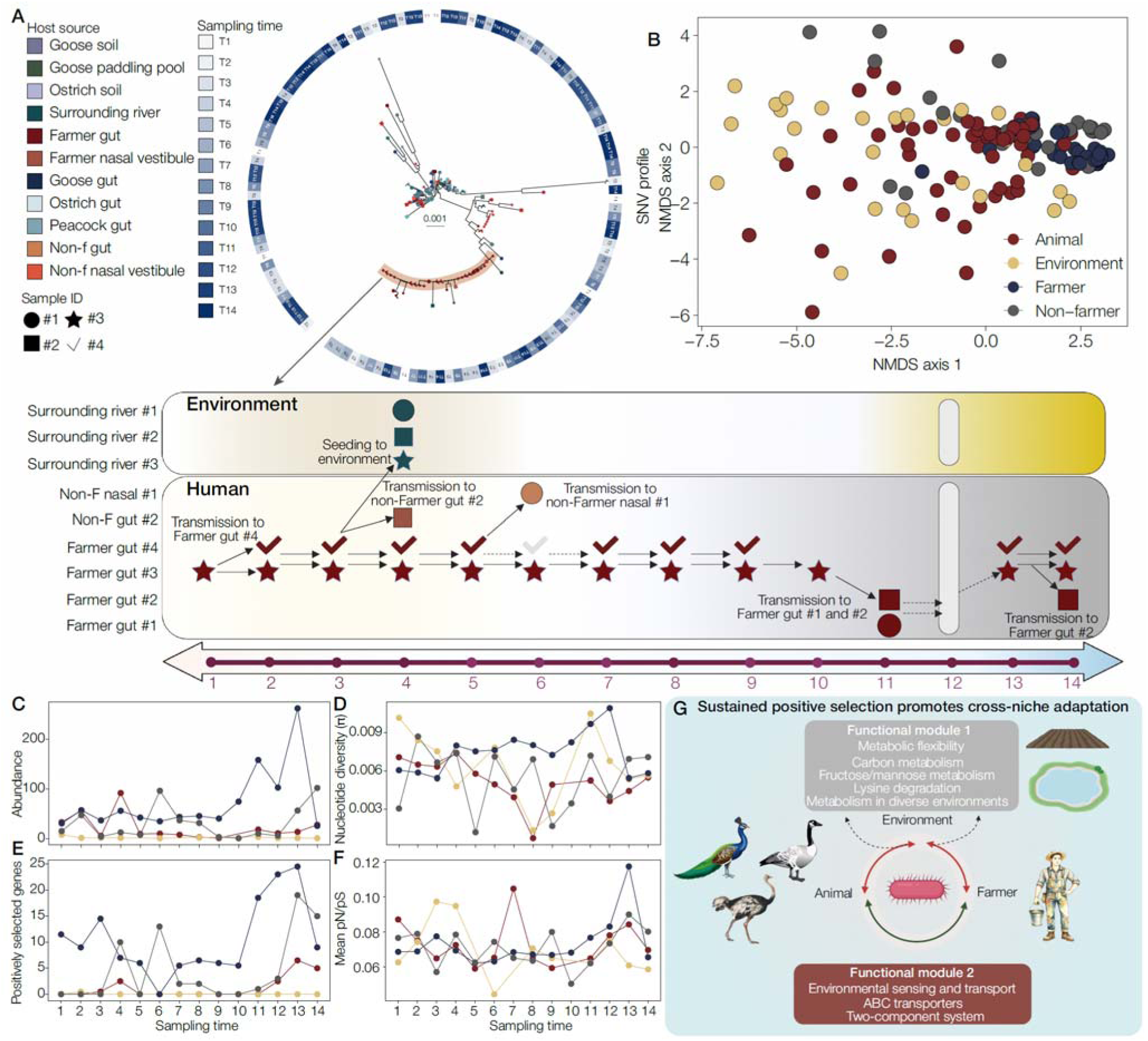
Adaptive dynamics accompany the cross-habitat transmission of a highly disseminated bacterial lineage. **A.** Strain-level profiling for E. coli GF12EGFS_2_bin.26 at 95% ANI. The upper panel shows the phylogenetic relationships among strain-resolved genomes recovered from the ecosystem, and the lower schematic summarizes one inferred transmission chain within the terminal cluster highlighted above. Sample identity is indicated by host source color and participant ID shape (#1: circle; #2: square; #3: star; #4 check mark). In the lower panel, each symbol represents a strain-positive sample at a given sampling time; grey bar at T12 indicates that the lineage was not detected or could not be typed at strain level. Arrows denote the most likely transmission links inferred from phylogenetic proximity and temporal continuity. **B.** NMDS ordination of SNV profiles for GF12EGFS_2_bin.26. Samples with more similar SNV compositions cluster more closely in ordination space, revealing partial grouping by habitat or host context. **C.** Absolute abundance dynamics of GF12EGFS_2_bin.26 across sampling timepoints, showing marked temporal fluctuation and a late increase in farmer-associated samples at the 13^th^ sampling. **D.** Temporal changes in nucleotide diversity of GF12EGFS_2_bin.26. Although diversity fluctuated over time, farmer-associated populations generally maintained relatively high nucleotide diversity, suggesting persistence of a genetically heterogeneous strain assemblage. **E.** Number of genes inferred to be under positive selection across sampling timepoints (McDonald–Kreitman two-sided F-statistic, P < 0.05). Positive selection intensified during later stages of dissemination. **F.** Mean pN/pS ratio over time, indicating shifts in selective pressure accompanying lineage turnover and transmission dynamics. **G.** KEGG-based functional summary of genes showing significant positive selection after sharing events. Enriched functions suggest adaptive changes associated with two modules. Detailed functional annotations are provided in **Table S3**.

Non-metric multidimensional scaling (NMDS) of single-nucleotide variant (SNV) profiles showed that samples recovered from environmental compartments occupied a broader and more dispersed genetic space, whereas those from farmer- and animal-associated niches were more constrained (**Fig. 5B**). Across the sampling period, this SGB remained detectable in multiple compartments and became particularly abundant in farmers during the later phase of sampling (**Fig. 5C**). Changes in strain composition coincided with corresponding shifts in genome-wide nucleotide diversity (**Fig. 5D**). These patterns could, in principle, reflect either the introduction of a new strain or changes in the relative abundances of pre-existing strains. To make the distinction, we quantified the number of previously unobserved SNVs in the mapped reads of each sample. We did not observe pronounced spikes in newly detected SNVs during the period in which abundance and nucleotide diversity increased (**Supplementary Fig. S3**), suggesting that the step-like genetic shift was unlikely to be explained solely by the sudden appearance of a highly divergent lineage.

To determine whether this stepwise genetic change was associated with adaptive selection, we quantified the number of genes showing evidence of positive selection using the McDonald-Kreitman framework^32^. The observed shift in strain composition coincided with a marked increase in the number of positively selected genes, consistent with selection acting on variants associated with expanding phylotypes. As the relative abundances of some strains increased, alleles specific to them appeared to undergo partial or soft selective sweeps (**Fig. 5E**). If strain composition had simply re-equilibrated after transmission, this signal would be expected to decay rapidly. Instead, the increase in positively selected genes persisted and was accompanied by an elevation in mean pN/pS ratios (**Fig. 5F**), suggesting continued turnover and selective filtering after host transition rather than passive stabilization of a newly established population. Having identified this selective signal, we examined which genes were repeatedly driven under positive selection during host-sharing events (see **Methods**). Functional prediction identified nine candidate genes associated with phylotype shifts during cross-compartment sharing, which grouped into distinct functional modules depending on the interface involved (**Table S3**). During environment-to-animal and environment-to-farmer transitions, the dominant signal involved functions related to metabolic flexibility, including carbon metabolism, fructose/mannose metabolism, lysine degradation, and metabolism in diverse environments. In contrast, animal-to-farmer transitions were more strongly associated with genes involved in environmental sensing and transport, particularly ABC transporters and two-component regulatory systems (**Fig. 5G**).

Together, these observations suggest that successful cross-host spread of this lineage may be accompanied by within-lineage genetic turnover consistent with adaptation to new host and habitat contexts. Rather than simple passive persistence, the data support a model in which entry into a new niche is followed by changes in lineage composition and selective filtering, potentially increasing the likelihood of persistence and subsequent spread across the wider ecosystem (**Fig. 5G**). These patterns are consistent with coordinated shifts in functions related to metabolic flexibility, environmental sensing, transport, and host-associated ecological interactions.

### *K. pneumoniae* acts as an ecological generalist

Because the animal hosts sampled in this study included rare and exotic birds, whose roles in the dissemination of potential human pathogens remain poorly understood, we profiled the clinically important opportunistic pathogens in this ecosystem^33^. We classified bacterial species detected in this study as candidate human pathogens according to three criteria: (1) ESKAPE pathogens; (2) pathogens listed by WHO; and (3) pathogens not included in WHO priority lists but widely reported in the literature as potential zoonotic or human-associated agents (**Extended Fig. 10A; Table S4**). We identified 23 candidate pathogens among the 6,075 SGBs, including 3 ESKAPE pathogens, 11 human pathogens, and 9 zoonotic pathogens (**Table S5**). The most prevalent were the zoonotic foodborne pathogen *Aliarcobacter cryaerophilus* (n = 53, prevalence = 10.6%) and the ESKAPE pathogen *K. pneumoniae* (n = 33, prevalence = 6.6%). We therefore examined the temporal dynamics of the five most prevalent pathogens together with all detected ESKAPE pathogens because of their potential public health relevance (**Extended Fig. 10A**).

Although recovery rates varied among taxa and sample types, most pathogens, once detected in a given subject or habitat, were observed again at two or more time points, indicating that they were not restricted to single sporadic detections (**Extended Fig. 10B–I**). Combining ANI- and phylogeny-based analyses, we observed several distinct patterns. First, highly similar (≥99.95%) clusters of *Bacteroides fragilis* included genomes from both farmers and non-farmers, whereas *Staphylococcus aureus* was restricted to a single non-farmer nasal site (**Fig. 6B**). Second, the foodborne pathogen *A. cryaerophilus* was frequently detected in the guts of exotic birds, particularly geese and ostriches, as well as in their surrounding habitats (**Extended Fig. 11E**). Third, *A. baumannii* was detected in goose gut and across all environmental sample types early in the sampling period, but was not recovered at later time points. Among the detected ESKAPE pathogens, *K. pneumoniae* was the only species repeatedly observed across peacocks, geese, farmers, and environmental samples, whereas the other ESKAPE pathogens showed much narrower ecological distributions. This unusually broad distribution prompted us to test whether *K. pneumoniae* also showed stronger evidence of cross-compartment sharing than the other detected ESKAPE pathogens. To validate, we complemented MAG-based detection with culture-based isolation and successfully recovered cultured *K. pneumoniae* isolates from 84.85% (28/33) of MAG-positive events across sampling waves (**Fig. 6C; Table S6**).

**Figure 6.**
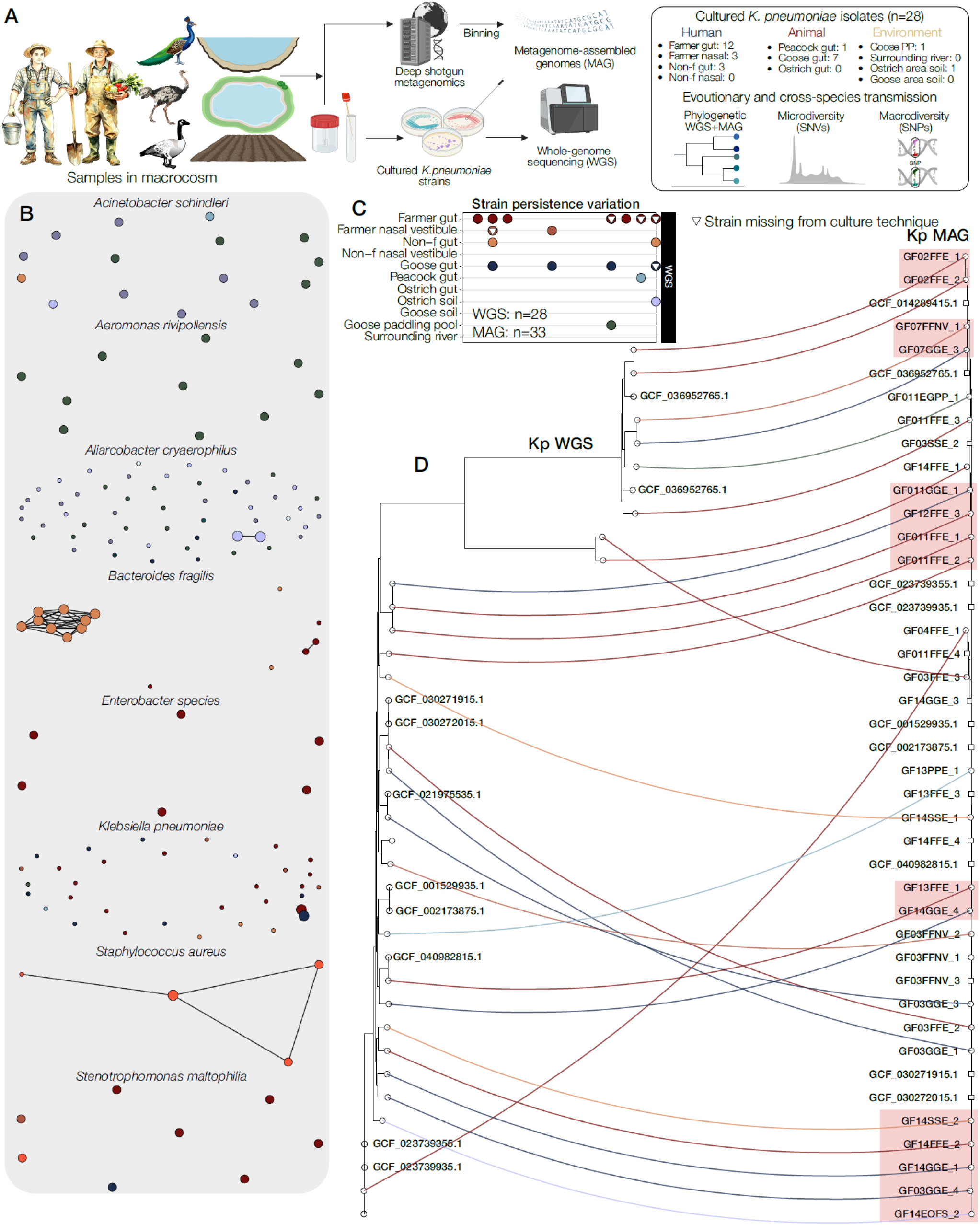
Zoonotic transmission and epidemiology of K. pneumoniae and other pathogens in the ecosystem. **A**. Overview of the analytical pipeline used to recover K. pneumoniae genomes from the ecosystem by combining isolate WGS and shotgun metagenomics. Samples were collected from human, animal and environmental compartments, followed by deep shotgun metagenomic sequencing, metagenome assembly and binning to reconstruct MAGs, together with culture-based isolation and WGS of K. pneumoniae strains. The table on the right summarizes the number of clinical K. pneumoniae isolates recovered from each sample type. **B**. Network analysis of pairwise ANI among MAGs assigned to major bacterial species recovered from the ecosystem. Each node represents one MAG, with node size proportional to the inferred number of sharing events. Edges indicate pairwise ANI values ≥99.96%. Edge lengths are shown for visualization only and are not quantitative. Node colors indicate distinct host or sample identities. **C**. Temporal recovery dynamics of K. pneumoniae strains inferred by WGS and MAG approaches. Colored points indicate recovery of K. pneumoniae from the corresponding host or habitat at each sampling timepoint. White triangles denote samples in which a K. pneumoniae MAG was recovered from metagenomic data but culture-based isolation failed (n = 6). Because multiple strain-positive samples could occur within the same sample category and timepoint, the number of plotted symbols may be lower than the true underlying strain count for that category at that time. A complete dot plot is shown in **Extended** Fig. 10. **D**. Tanglegram showing phylogenetic concordance between cultured K. pneumoniae isolates recovered by WGS (left) and K. pneumoniae MAGs reconstructed from shotgun metagenomes (right). The WGS phylogeny was inferred through maximum-likelihood inference using IQ-TREE, whereas the MAG phylogeny was reconstructed with GToTree based on amino acid alignments of 172 single-copy marker genes. To enable direct topological comparison, both trees were reconstructed by the same set of publicly available reference genomes representing the sequence types (STs) identified in the facility. Colored lines connect corresponding genomes across the two phylogenies. Tip shapes indicate genome origin (circles, genomes from this study; squares, reference genomes). Putative cross-compartment sharing events supported by both phylogenies are highlighted with red boxes.

We reconstructed *K. pneumoniae* phylogenies independently from cultured isolates and MAGs and compared them using a tanglegram (see **Methods**). Concordance between the two phylogenies supported the broad ecological distribution of *K. pneumoniae* and revealed at least five putative cross-compartment sharing chains, spanning hosts and habitats (**Fig. 6D**). In addition, closely related or near-identical strains were repeatedly detected across multiple hosts and habitats, consistent with repeated sharing events within the ecosystem. Although the precise sources and directionality of movement cannot be resolved from these data, the repeated detection of near-identical strains across compartments supports sustained cross-host circulation of K. pneumoniae within this ecosystem. Together, these findings distinguish *K. pneumoniae* from the other detected ESKAPE pathogens by its unusually broad ecological range and repeated occurrence across human, animal, and environmental compartments (**Fig. 6C-D**).

## Discussion

Understanding how microbiomes and antimicrobial resistance genes (ARGs) circulate across human-animal-environment interfaces is still a central One Health challenge^13^. Using a strain-resolved metagenomic framework encompassing 6,075 SGBs, we provide quantitative evidence that animal-associated occupancy reshapes the microbiome and resistome of surrounding environments and occupationally exposed farmers through repeated contact. We further show, by tracking a frequently shared lineage, that cross-host spread is accompanied by within-lineage genetic turnover and selection-consistent changes, suggestive of ecological filtering during movement across hosts and habitats. Finally, by integrating longitudinal strain-resolved metagenomics with isolate-based whole-genome sequencing, our results support a previously underappreciated circulation pattern for *K. pneumoniae*, raising the concern that its ecological versatility would facilitate the acquisition and dissemination of both virulence determinants and mobile ARGs^19,34^.

A major strength of this study is the longitudinal strain-resolved design, which enabled us to distinguish sharing patterns across human, animal, and environmental compartments at finer resolution^3,5,35–37^. By comparing farmers with age- and village-matched non-farm residents, while simultaneously sampling nearby environmental and animal-associated habitats, we show that animal occupancy was associated with measurable restructuring of both microbiome composition and resistome architecture. In particular, animal-associated habitats emerged as the dominant interface for strain and ARG sharing, whereas direct sharing between humans and the broader surrounding environment was comparatively limited. Among human-associated links, animals appeared to be more important counterparts than environmental compartments. Together, these patterns support a model in which animals and their immediate habitats form the main ecological interface sustaining microbial and resistance sharing in this ecosystem. This interpretation is strengthened by the maintenance and sharing of clinically important ARGs, including *bla*_NDM_, *mcr*-family, and *vanYB* in the context of widespread MGE co-occurrence.

Beyond describing sharing patterns, our results also provide clues as to why only a subset of lineages persist and spread across compartments. Analysis of one widely shared lineage revealed within-lineage genetic turnover together with selection-consistent changes after entry into new host or habitat contexts. Previous microbiome studies have shown that strain-level microdiversity can strongly shape microbial dynamics, with closely related strains often occupying distinct ecological niches in systems such as anaerobic digesters^38^, oceans^39,40^, and geothermal habitats^41^. Strains have been described through both ecological frameworks, including metapopulation structure in subseafloor environments^42^ and carrying capacity in the human gut^43,44^, and evolutionary frameworks, such as modes of diversification in lake ecosystems^45,46^. In this context, our findings are consistent with the view that the patterns observed here reflect ecological filtering and selection acting on strain-level variation, plausibly driven by differences in nutrient availability and host-associated environments. Longer-term studies will be needed to determine how these dynamics interact with broader ecological processes over time^47^.

An unexpected finding of our study was the distinctive ecological behavior of *K. pneumoniae*^48,49^. By integrating MAG-based strain tracking with isolate-based whole-genome sequencing, we proved that *K. pneumoniae* was the only detected ESKAPE pathogen showing broad and repeated occurrence across animal, human, and environmental compartments. Repeated detection of *K. pneumoniae* carriage in multiple residents, together with its persistence across compartments, suggests an underappreciated circulation pattern at the human–animal–environment interface^34^. Given prior evidence that *K. pneumoniae* undergoes convergent evolution, including recurrent acquisition of virulence loci such as *iuc3*^50^, our results further raise the question that whether its broad ecological versatility may facilitate both ARG persistence and the acquisition of virulence-associated traits at the human–animal–environment interface^51,52^.

The ecological context of this study further strengthens the interpretation of the observed sharing patterns. The extensive animal occupancy and relative isolation of this rural ecosystem make repeated external introductions a less parsimonious explanation for the structured sharing observed here^11^. Moreover, the sampling design enabled us to distinguish likely direct from indirect ecological links within the system. Permutation-based analyses showed stronger sharing between geese and goose-associated soil than with ostrich-associated soil, between ostriches and ostrich-associated soil than with goose-associated soil, and between geese and their paddling pool than with the surrounding river. These compartment-specific associations strengthen the inference that transmission was structured primarily by within-ecosystem animal occupancy rather than diffuse background exposure.

Several limitations should be considered. First, both ARG dissemination and strain sharing were dynamic and strongly right-skewed over time. Although sharing intensified toward the end of sampling (T13), we lacked sufficient evidence to attribute this shift to specific ecological drivers. Second, because isolate-based sampling initially focused on *K. pneumoniae*, we did not systematically culture other ESKAPE pathogens. The apparent ecological distinctiveness of *K. pneumoniae* among ESKAPE therefore requires validation in future studies explicitly designed for comparative pathogen isolation. Even so, its dominance in MAG recovery and reproducible detection across independent analytical frameworks argue against this pattern being purely artefactual^12^. Finally, our longitudinal surveillance was conducted in a single ecosystem over one year, largely because of site accessibility and the ecological uniqueness of this farm. Although this setting provided a rare opportunity to resolve microbial and resistome dynamics in a relatively self-contained agricultural system, it may limit the generalizability of our findings. At the same time, its ecological characteristics enabled evaluation of key hypotheses under reduced confounding. Multi-site studies with longer follow-up will be needed to assess how broadly these patterns extend across ecological farming systems.

Overall, our findings show that farming-associated ecological exposure can durably reshape the microbiomes of farmers and their surrounding biotopes through strain transmission and ARG dynamics. We highlight the cryptic persistence and ecological generalist potential of opportunistic pathogens within connected agricultural ecosystems. Animals act as centrality in driving microbial circulation across human and environment in a connected ecosystem, with implications for functional change, pathogen ecology, and resistance dissemination. Future integration of long-read sequencing will be important for resolving the movement of phages, plasmids, operons, and other mobile genetic elements, while experimental approaches will be needed to identify the microbial traits that favor persistence under different ecological contexts^53,54^. Such efforts will help clarify how animal-associated exposure affects human and environmental microbiomes and how these processes intersect with the epidemiology of opportunistic pathogens and AMR^54^.

## Data Availability

Shotgun metagenomic data for OH-MicroDynamics are available at the NCBI SRA (PRJNA1457641), with respective sample-wise metadata information available in Table S1. Accession numbers for K. pneumonia WGS datasets are available at the NCBI SRA under accession number (PRJNA1456307 and PRJNA1456406).

https://www.ncbi.nlm.nih.gov/sra/?term=PRJNA1457641

https://www.ncbi.nlm.nih.gov/sra/?term=PRJNA1456307

https://www.ncbi.nlm.nih.gov/sra/?term=PRJNA1456406

## Acknowledgments

We sincerely thank all participants for their contributions to this year-long study. We are particularly grateful to Kaifeng Miaolin Agricultural Development Co., Ltd., and Ms. Miao Wang for their generous and sustained support, including assistance with sample collection, preservation, transportation, and field training. This work would not have been possible without their continued commitment. H.W. acknowledges financial support from the Henan Province Key Research and Development Project (Key Project of International Science and Technology Cooperation of Henan Province; grant agreement no. 241111521100) and the Henan Province Science and Technology Research Project (grant agreement no. 252102111013). J.X. acknowledges support from the Young Postgraduate Fund of the National Natural Science Foundation of China (NSFC; grant agreement no. 824B2101), the Young Elite Scientists Sponsorship Program of the China Association for Science and Technology (CAST), and the Fudan University Keqing–Zhenhua Development Fund (Young Scholar Program in Global Health). G.B. acknowledges support from the Research Foundation - Flanders (“Fonds voor Wetenschappelijk Onderzoek - Vlaanderen,” G098321N), from the European Union Horizon 2023 RIA project LEAPS (grant agreement no. 101094685), and from the DURABLE EU4Health project 02/2023-01/2027, which is co-funded by the European Union (call EU4H-2021-PJ4) under grant agreement no. 101102733. C.L. acknowledges support from the Hong Kong RGC Junior Research Fellow Scheme (JRFS2526-5S05). A.W.D acknowledges support from NIH/NIAID 5T32AI155391-05. Z.J acknowledges support from Key Public Health Discipline Foundation of Minhang District (grant agreement no. MGWXK2023-01) and the construction plan of key disciplines in Shanghai Municipal Health System [grant agreement no. 2024ZDXK0017].

## Author contributions

Conceptualization: J.X., Z.X., T.C., and H.W.

Sample collection and data curation: Y.Z., H.Z., and J.Z. (Junkai Zhang).

Methodology: J.X., Z.X., and H.W.

Visualization: J.X., Z.X., L.Z., and W.G.

Validation: J.L. (Jianhua Liu), Y.P., T.C., and H.W.

Writing – original draft: J.X. and Z.X.

Writing – review & editing: J.L. (Jian Li), G.B., A.W.D., C.L., J.Z. (Jinxin Zhao), T.C., and H.W.

Funding acquisition: J.L. (Jian Li), H.W. and T.C.

Supervision: J.L. (Jian Li), J.Z. (Jinxin Zhao), T.C., and H.W.

All authors read and approved the final manuscript.

## Competing interests

All authors declared no conflict of interests.

## Data availability

Shotgun metagenomic data for OH-MicroDynamics are available at the NCBI SRA (PRJNA1457641), with respective sample-wise metadata information available in Table S1. Accession numbers for *K. pneumonia* WGS datasets are available at the NCBI SRA under accession number (PRJNA1456307 and PRJNA1456406).

## Code availability

All statistical analyses were performed using CRAN R (v4.5.0) and Python (v3.11). Codes are deposited into GitHub: https://github.com/jobsxing/Eco_microbiome

## Methods

### Ethics statement

All study procedures complied with relevant institutional and ethical regulations. Sampling protocols were reviewed and approved by the Scientific Ethics Committee of Henan Agricultural University (HENAU-SEC; approval ID: HNND2024110101). All biological specimens were collected under HENAU-SEC authorization. Written informed consent was obtained from all participants prior to enrollment. Both farmers and non-farming participants received financial compensation for their participation.

### Study design and rationale

Antibiotic use in livestock production has been increasingly restricted in China through a series of regulatory actions, including the removal of antibiotic feed additives from animal husbandry in 2021. Despite these efforts, the longitudinal ecological dynamics of microbiomes and resistomes in antibiotic-free farming systems remain poorly understood.

To address this gap, we identified a small ecological farm (hereafter referred to as the “ecosystem”) in Xijugang Village, Henan Province, China (114.4°E, 34.4°N), where ostriches (*Struthio camelus*), peacocks (*Pavo cristatus*) and geese (*Anser cygnoides domesticus*) were raised under free-ranging, low-intensity management conditions (**Extended Fig. 1**). Animals were maintained on plant-based diets consisting of alfalfa, maize mash and soybean meal. No antibiotics had been used on the farm since 2022, as confirmed through farm management records and continuous field monitoring during the year preceding sample collection. We briefly described the study site in the **Supplementary Note 1**.

Using this antibiotic-free ecological setting as a natural One Health study system, we established a prospective longitudinal cohort integrating human, animal and environmental sampling. The study was designed to characterize the temporal dynamics, cross-compartment sharing patterns and ecological convergence of microbial communities and resistance determinants across the human–animal–environment interface in the absence of routine antibiotic use.

### Participant recruitment

At the ecosystem, four full-time workers were initially screened for eligibility. To minimize potential confounding related to recent antimicrobial exposure, all farm workers were confirmed to have had no antibiotic exposure within the preceding six months. Eligible farm workers were 30–40 years of age. To establish a demographically comparable control group, three non-farm residents from the same village were recruited and matched to farm workers by age and sex. Non-farm participants were also confirmed to have had no antibiotic exposure within the preceding six months and were 30–40 years of age.

Recruiting non-farm residents from the same village helped ensure broadly comparable cultural, dietary and socioeconomic backgrounds, thereby reducing potential environmental and lifestyle confounding. Farm-worker eligibility criteria included full-time employment at the ecosystem (>8 h per day), age >18 years, and continuous employment for at least one year before recruitment. Non-farm participants were >18 years of age and had neither lived nor worked on any farm during the preceding three years.

All participants provided written informed consent prior to enrollment. In addition, all volunteers completed a metadata questionnaire at study entry. The questionnaire collected information on date of birth, sex, anthropometric measurements (body weight and height), medical care received within the year preceding study initiation, and chronic health conditions. Information on household composition (i.e., permanent household members) was also collected on an optional basis. The questionnaire further confirmed that none of the enrolled participants had been exposed to or had consumed any probiotic products before enrollment. All participants were enrolled for longitudinal follow-up and provided repeated samples over the course of the study. Compensation was provided upon enrollment.

### Sample collection and processing

A year-long longitudinal sampling campaign was conducted to characterize microbiome and resistome dynamics across the human-animal-environment interface. Samples were collected from three compartments: humans, animals, and the environment. The sampling schedule consisted of two phases: Intensive phase: sampling every 10 days during the first 3 months; Routine phase: sampling every 2 months during the subsequent 9 months. All metadata are provided in **Table S1**.

#### Human samples

For human participants, both nasal vestibule and fecal samples were collected at each sampling time point.

Nasal samples were collected using DNA/RNA Shield Collection Tubes with Swabs (R1106; Zymo Research). A sterile swab was inserted into each anterior nare and gently rotated to collect nasal microbiota. Swabs from both nostrils were pooled to maximize biomass and reduce intra-individual variability. All nasal samples were collected by **trained study personnel** following standardized protocols to ensure procedural consistency.

For fecal sampling, given the sensitive nature of fecal sample collection, participants received standardized training prior to study initiation to ensure consistent self-collection. Each participant was provided with a DNA/RNA Shield Fecal Collection Tube (R1101; Zymo Research). Participants deposited fresh stool onto sterile collection paper and transferred a small aliquot into the collection tube using the provided swab. Samples were temporarily stored at −20 °C and retrieved by field investigators within 24 h. All samples were handled using identical protocols to minimize potential bias associated with storage conditions.

#### Animal samples

Fresh fecal samples were collected from three livestock species maintained within the ecosystem: geese (*Anser cygnoides domesticus*), ostriches (*Struthio camelus*) and peacocks (*Pavo cristatus*). For each species, four ID-code-matched individuals were selected and monitored longitudinally throughout the study period. These animals were maintained within the same farming unit and experienced comparable environmental exposures. Fresh fecal samples from each individual were collected at every sampling time point.

#### Environmental samples

Environmental samples were collected concurrently with human and animal sampling and included water and soil associated with animal habitats.

Water samples were obtained from two sources: (i) a goose paddling pool and (ii) surrounding river water. At each time point, four spatially distinct subsamples (500 mL each) were collected per water source. For river sampling, subsamples were obtained at varying depths (surface to ∼10–20 cm below surface) to capture vertical heterogeneity. For the goose paddling water, subsamples were collected from different locations and depths within the water column. Subsamples from each source were combined to generate one composite sample per habitat per time point, thereby integrating spatial and depth-related variability. Water samples were filtered through 0.22 μm polycarbonate membranes (Millipore, USA). The membranes were aseptically transferred to sterile petri dishes, cut into fragments and transferred into sterile 2 mL centrifuge tubes within a biosafety cabinet.

Soil samples were collected from feeding areas within goose and ostrich habitats. At each time point, multiple subsamples were collected from distinct locations within each habitat and pooled to generate composite samples. Care was taken to avoid areas with visible fecal contamination, and sampling was conducted beneath the immediate surface layer to minimize direct fecal input while retaining habitat-associated microbial signatures.

#### Sample transport and storage

Across the entire study period, 500 biological samples were collected from human, animal and environmental compartments (sample composition summarized in **Fig. 1A**). The overall sampling workflow and longitudinal timeline are illustrated in **Fig. 1A-B**. All samples were temporarily stored at -20 °C and transported on ice to Henan Agricultural University within 48h of collection and stored at -80°C until DNA extraction.

### DNA extraction and quantification

Approximately 100 mg of frozen fecal material was processed under a biosafety cabinet using sterile spatulas. Gloves were changed and the workspace disinfected with 10% bleach between samples. Metagenomic DNA was extracted using the QIAamp DNA Microbiome Kit (QIAGEN, USA) for both fecal and nasal specimens (the latter thawed on ice before extraction). Each nasal sample combined two biological replicates to enhance DNA yield. DNA concentrations were measured with a Qubit fluorometer (Thermo Fisher Scientific, USA) using dsDNA assays. Negative extraction controls (ddH_2_O) were included in each batch; only uncontaminated batches proceeded to library construction. Note that the peacock samples from the first sampling event were suspected contaminated and excluded for subsequent analysis.

### Deep metagenomic sequencing

For each fecal sample, the extracted DNA was diluted to 0.5 ng/μL with a total of 20 ng and subsequently used as the input for Illumina sequencing library preparation using the Nextera kit (Illumina, USA). The libraries were purified using AMPure XP beads, pooled, and sequenced on the NovaSeq 6000 platform (Illumina) to obtain 2 × 150-bp paired-end reads. Each sample was ensured to have a minimum sequencing depth of 15 Gb to guarantee the robustness of sequencing depth. Adapters and low-quality reads were removed using Trimmomatic v0.38 (ILLUMINACLIP:NexteraPE-PE.fa:2:30:10:1:true, SLIDINGWINDOW:4:20, LEADING:10, TRAILING:10, MINLEN:60)^55^. Host-derived reads were filtered out with DeconSeq v4.3^56^, and unpaired reads were excluded using repair.sh from BBMap v40.02. The resulting reads were retained as clean reads for downstream metagenomic analyses.

### Microbiome profiling at species-level

Taxonomic profiles were generated using MetaPhlAn v4 with default parameters and an abundance threshold of 0.1%^57^. Across the 500 samples, this yielded 28,189 species-level features, 9,882 genus-level taxa, and 3,080 family-level taxa. Functional annotation was performed using HUMAnN3 on the same reads to infer gene-family and pathway-level functional repertoires^24^.

### Resistome profiling

ARGs were identified using ARGs-OAP v3.2.2 by aligning quality-filtered metagenomic reads against the SARG v3.0_S database^26^. Alignments were retained using the following thresholds: ≥80% sequence identity, ≥75% alignment coverage, and an e-value ≤1 × 10^-7^. ARGs were hierarchically classified into type, subtype, and gene (reference sequence) levels based on the SARG annotation framework^26^. To enable cross-sample comparisons, ARG abundances were normalized to copies per cell, calculated using the abundance of single-copy marker genes, which approximates copies per genome under the assumption of one genome per cell. This normalization accounts for sequencing depth, gene length and microbial genome content, thereby providing a robust estimate of relative ARG abundance across samples. To further assess the potential public health relevance of detected resistance determinants, ARGs identified at the gene level were additionally annotated according to the ARG risk-ranking framework provided in the SARG v3.0_S database^28^.

### Benchmarking of metagenome assembly performance

To identify an optimal genome reconstruction strategy across heterogeneous habitats, we benchmarked three commonly used binning paradigms: co-assembly, single-sample binning and multi-sample binning^58^. Co-assembly jointly assembles reads from all samples and bins contigs based on cross-sample coverage profiles, leveraging co-abundance information but potentially introducing chimerism and obscuring sample-specific variation. In contrast, single-sample binning performs assembly and binning independently for each sample, preserving within-sample heterogeneity but ignoring cross-sample abundance patterns. Multi-sample binning combines individual assemblies with cross-sample coverage information through all-against-all comparisons, often yielding more complete and higher-quality metagenome-assembled genomes (MAGs), albeit at increased computational cost.

Given that binning performance is known to vary across sample types, we systematically evaluated these strategies across 11 habitat types spanning human, animal and environmental compartments, including non-farmer nasal vestibule, non-farmer gut, farmer nasal vestibule, farmer gut, ostrich gut, goose gut, ostrich soil, goose soil, goose paddling pool, and surrounding river^58^. For each habitat, 10 temporally distinct samples were randomly selected to minimize sampling bias.

Reads were assembled using MEGAHIT v1.2.9^59^, and contigs were binned using CONCOCT v1.1.0^60^, MaxBin 2 v2.2.7^61^, and MetaBAT 2 v2.15^62^. Bin sets generated by all tools were subsequently reconciled and refined using MetaWRAP v1.3.2 to produce a final high-quality MAG catalogue.

Benchmarking results are provided in the **Supplementary Note 2** and were used to guide the selection of assembly and binning strategies for downstream analyses.

### MAG recovery

Given the demonstrated superiority of the multi-sample binning strategy, this approach was used for subsequent reconstruction of MAG profiles. To account for substantial microbial heterogeneity across habitats, each habitat type was treated as an independent sample group. Accordingly, all-against-all coverage comparisons were performed within each habitat group, with coverage calculated by pooling samples from the same habitat, resulting in 29,838 filtered MAGs. Bin quality - including completeness and contamination - was assessed using CheckM2 v0.1.3^63^, and taxonomy was assigned using the Genome Taxonomy Database Toolkit (GTDB-Tk v2.1.1, GTDB release 207)^64^. A total of 29,838 genome bins with ≥50% completeness and <10% contamination were retained for downstream analysis and dereplicated at 96% average nucleotide identity (ANI) using dRep v3.4.0^65^. To select an appropriate dereplication threshold, we ran dRep across ANI cutoffs ranging from 90% to 99% and evaluated both the resulting number of dereplicated genome groups and the extent to which bins originating from the same assembly were merged. We selected 96% ANI because it was located immediately before a sharp increase in the total number of dereplicated genome groups, while minimizing erroneous merging of bins from the same assembly; specifically, only one of the 29,838 retained bins was grouped with another bin derived from the same assembly at this threshold. This cutoff therefore provided a practical balance between maximizing species-level resolution and avoiding competitive read mapping among nearly identical strains. Applying the 96% ANI threshold resulted in 6,075 representative bacterial species-level genome bins (SGBs), which were treated as species-level units throughout this study. Details of these unique SGBs are provided in **Table S2**. For phylogenetic reconstruction, GTDB-Tk identified 120 and 122 universal marker genes for bacterial and archaeal genomes, respectively, and generated concatenated multiple-sequence alignments of the predicted marker genes for each representative genome. Based on these alignments, a maximum-likelihood phylogenetic tree was inferred using IQ-TREE v2.2^66^ , with the best-fit substitution model selected by ModelFinder and branch support assessed using 1,000 ultrafast bootstrap (UFBOOT) replicates. The resulting tree was visualized with iTOL v7^67^.

### Sharing dynamics of ARG, mobile genetic elements and horizontal genes transfer

To characterize the mobilization and dissemination potential of ARGs across our ecosystem, we conducted two complementary analyses. First, we identified ARGs and mobile genetic elements (MGEs) within MAGs and evaluated the dynamics of ARGs potentially associated with horizontal mobility. Second, we quantified horizontal gene transfer (HGT) events among the three ecological compartments of the study system-farmers, animals and the environment.

#### Annotation of ARGs, MGEs and genetic context

Open reading frames (ORFs) were predicted from assembled contigs using Prodigal v2.6.3 with the “-p meta” parameter. Quality-filtered reads were mapped back to predicted ORFs using BWA v0.7.17^68^, and gene abundance was quantified as reads per kilobase per million mapped reads (RPKM) using SAMtools v1.18^69^ and CoverM v0.6.1^70^. To normalize gene abundance across samples, the abundance of each ORF was divided by the mean RPKM of 40 universal single-copy bacterial marker genes, thereby estimating gene abundance in units of copies per genome equivalent^71^. These marker genes were identified using the FetchMG pipeline (available at https://github.com/motu-tool/FetchMGs/tree/main) with 40 Hidden Markov Models (HMMs)^72^ and detected using HMMER v3.4 with an e-value threshold of 1 × 10^-10^. Predicted ORFs were subsequently aligned against the ResFinder database^73^ using DIAMOND BLASTX v2.15.0 to identify ARGs. MGEs were detected by aligning contigs against the TnCentral^74^, ISfinder^75^, and INTEGRALL^76^ databases using BLASTN v2.15.0. The datasets used for these reference databases corresponded to the most recent releases retrieved as of December 2025.

ARG annotations were retained if alignments met the thresholds of ≥80% amino acid identity and ≥70% alignment coverage relative to the reference sequence. MGE annotations were retained using thresholds of ≥80% nucleotide identity and ≥50% alignment coverage^33^. Identified ARGs and MGEs were grouped according to antibiotic resistance classes and MGE categories defined in their respective databases. Because different ARG databases may use inconsistent subtype naming conventions, we implemented an additional filtering step prior to ARG annotation. Specifically, ARG sequences in the ResFinder database were retained only if they exhibited 100% nucleotide identity and 100% alignment coverage with corresponding entries in either the CARD or AR gene catalogue. This strategy ensured consistent subtype nomenclature across databases and improved the reliability of downstream analyses, although it may exclude certain ARG variants. Contigs were further classified as plasmid-derived using PlasFlow (v1.1.0)^77^ with a probability threshold of 0.9 and a minimum contig length of 2 kb. To evaluate potential ARG mobilization, co-occurrence between ARGs and MGEs was defined when both elements were located on the same contig within a 10 kb genomic distance, either upstream or downstream, consistent with criteria used in previous analyses of mammalian microbiomes^33^.

#### Dynamics of horizontal gene transfer

HGT events were identified following previously established approaches for detecting recent gene transfer among microbial genomes^78^. Contigs from high- and medium-quality SGBs reconstructed from animal samples were aligned against SGBs from farmer, non-farmer, and environmental samples using BLASTN (v2.15.0) in an all-versus-all comparison. Pairwise ANI between genome bins was calculated using FastANI (v1.33)^79^. HGT events were defined as genomic fragments >500 bp in length that were shared between MAG pairs with >99.5% nucleotide identity, while the overall genome similarity between the two MAGs remained <95% ANI or they were classified as distinct species according to GTDB-Tk taxonomy^3,27^. To further assess the contribution of resistance gene transfer, each detected HGT fragment was cross-referenced with ARG annotations identified in the previous step. HGT events were subsequently categorized as ARG-associated HGT (ARG-HGT) if the transferred genomic fragment contained an ARG, and as non-ARG HGT otherwise.

### Microbial strain sharing and ecological transmission analysis

Single-nucleotide variants (SNVs) and microbial lineage sharing across shotgun metagenomes were inferred using inStrain (v1.5.7)^80^. To enable strain-resolved comparisons, all dereplicated MAGs were concatenated into a single FASTA file to generate a reference genome catalogue comprising 6,075 representative species-level genome bins (SGBs) (see MAG recovery section). To reduce the computational burden associated with exhaustive pairwise comparisons across ∼500 shotgun metagenomes, we generated a reduced candidate genome catalogue enriched for potentially shared lineages. Specifically, the representative SGBs were further clustered using dRep v3.2.2 (-sa 0.98, -nc 0.1), yielding 3,987 candidate genome clusters for downstream comparison.

Quality-filtered metagenomic reads from all samples were then profiled against this reduced catalogue using the inStrain *profile* module to quantify population-level genetic variation and genome coverage for each SGB. An SGB was considered confidently detected if it exhibited genomic breadth ≥50% in a sample, resulting in 2,571 SGBs retained for strain-level comparisons. Pairwise genome comparisons were subsequently performed using inStrain *compare*, restricted to sample pairs in which the corresponding genome met the detection threshold. For each sample pair, population average nucleotide identity (popANI) and the fraction of the genome compared (percent_genome_compared) were calculated. Genome pairs with popANI ≥99.999% and percent_genome_compared ≥50% were classified as strain-sharing events, consistent with the presence of near-identical microbial lineages in both samples, following previously described criteria^3^.

All human and animal samples were linked to unique subject identifiers, and all environmental samples were linked to unique sample identifiers and collection time points. This enabled pairwise quantification of strain sharing across all eligible sample pairs within each sampling round. Temporal dynamics of strain sharing were evaluated by restricting comparisons to samples collected at the same sampling time point. For each sampling round, we quantified the number of shared strains for every eligible sample pair and summarized these events across major habitat interfaces, including farmer–animal, farmer–environment, non-farmer–animal, non-farmer–environment, and animal–environment comparisons (**Fig. 4D-E**). To illustrate how lineage-sharing patterns varied across specific sample types over time, we additionally summarized strain-sharing events at the first (T01) and last (T14) sampling rounds at the level of individual sample-type combinations (**Fig. 4D**). Differences in lineage-sharing patterns between occupational groups were assessed using negative binomial regression, in which each observation corresponded to one eligible, time-matched sample pair and the response variable was the number of shared strains detected in that pair. Fixed effects included human type (farmer versus non-farmer), interface type, and their interaction (**Fig. 4F**). Additional permutation-based analyses were performed to test whether strain sharing between specific gut habitats and environmental reservoirs exceeded random expectations under both global and time-stratified null models (see Statistical analyses section; **Extended Fig. 9C**).

To investigate how strain-sharing events were distributed across the ecosystem, pairwise comparison results were integrated with metadata including habitat type, host category, sampling round, and subject or sample identifiers. Samples were assigned to four major ecological groups: farmer, non-farmer, animal, and environment. Environmental samples included goose soil, ostrich soil, surrounding river water, and goose paddling pool; animal samples included goose, ostrich, and peacock gut samples; and human samples comprised gut and nasal vestibule samples from farmers and non-farmers. For each habitat pair, sharing intensity was calculated as the number of observed strain-sharing events divided by the total number of eligible sample-pair combinations for that habitat pair, yielding an opportunity-adjusted estimate of lineage sharing (**Fig. 4G**).

To further characterize the ecological breadth of reconstructed genomes, relative abundance profiles were integrated with metadata to determine habitat occupancy (**Fig. 4C**). Genome presence was evaluated using both lenient and stringent definitions. Unless otherwise stated, all presence/absence-based analyses were based on the stringent definition. Under this definition, a genome was considered present in a given sample only if its relative abundance exceeded the larger of (i) the 10th percentile of all non-zero genome relative abundances within that sample and (ii) a global abundance floor of 1 × 10^-5^. This thresholding scheme was used to reduce spurious presence calls driven by extremely low-abundance mappings. Based on this definition, genomes were classified according to their occupancy across the four ecological groups, enabling quantification of cross-habitat spillover and ecological niche breadth using Shannon entropy^81^ (**Fig. 4C**).

Habitat similarity in genome composition was assessed using the Jaccard index:

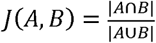

where *A* and *H* represent the sets of detected genomes in the two habitats being compared (**Fig. 4H**).

We next summarized strain-sharing events at the taxonomic level to evaluate whether sharing was broadly distributed across taxa or concentrated within a limited subset of lineages (**Fig. 4I-J**). For each habitat interface, shared genomes were taxonomically annotated and summarized as counts of strain-sharing events per taxon. Bray-Curtis dissimilarity was then calculated on these taxonomic count profiles, followed by Principal Coordinate Analysis (PCoA) (**Fig. 4J**). Sharing events were further aggregated across all cross-habitat comparisons, and concentration was quantified using Lorenz-like distributions^30^ and Gini coefficients^82^, enabling identification of genomes that disproportionately contributed to lineage sharing (**Fig. 4K**).

To summarize the ecosystem-wide dissemination potential of individual genomes, we defined a cross-habitat connectivity index (CCI) integrating four components: habitat niche breadth, number of habitats occupied, contribution to lineage-sharing events, and prevalence in human-associated samples. After scaling each component to the range 0–1, the composite score was calculated as:

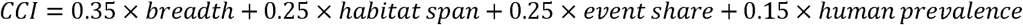

Weights were assigned a priori to place slightly greater emphasis on ecological breadth while retaining substantial contributions from habitat span, sharing frequency, and prevalence in human-associated samples. Genomes with the highest CCI values were interpreted as candidate genomes with high cross-habitat dissemination potential within the ecosystem (**Fig. 4L; Extended Fig. 9B**).

### Genome-scale evolutionary dynamics of a highly disseminated lineage

To investigate genome-scale evolutionary dynamics following cross-habitat dissemination, we selected the representative genome with the highest strain-sharing frequency as the focal lineage for downstream strain-resolved analyses. Genome-wide coverage, breadth, nucleotide diversity, SNV profiles, and gene-level evolutionary statistics were extracted from inStrain outputs. Samples with insufficient genome recovery were excluded, and genome-wide diversity analyses were restricted to samples with coverage ≥5 and breadth ≥0.5.

To place the focal lineage in phylogenetic context, we reconstructed a phylogenomic tree for the corresponding dRep-defined cluster (ANI ≥96%) using GToTree ^83^. Conserved orthologues were identified using the bacteria-specific HMM set comprising 74 single-copy core genes, followed by automated alignment and trimming. A maximum-likelihood phylogeny was then inferred using FastTree with the JTT amino acid substitution model and CAT approximation for site-specific evolutionary rates^84^. This phylogeny was used to contextualize the focal lineage among its closely related genomes.

To characterize genome-wide mutational structure across ecological compartments, non-metric multidimensional scaling (NMDS) was performed on SNV profiles of the focal lineage. Within-sample evolutionary dynamics were summarized using complementary genome- and gene-level metrics. Nucleotide diversity (π) was used to quantify intrapopulation sequence diversity. To summarize shifts in selective constraint across habitats, we calculated sample-level mean pN/ps by averaging inStrain-derived gene-level pN/ps estimates across genes with valid values. In parallel, genes with pN/ps >1 and at least two nonsynonymous SNVs were designated as candidate genes with elevated nonsynonymous enrichment, and their numbers were quantified for each sample and time point. Temporal trajectories of lineage abundance, nucleotide diversity, mean pN/ps, and candidate genes with elevated nonsynonymous enrichment were then summarized across ecological compartments and longitudinal sampling schemes.

To identify genes showing recurrent evidence of adaptive evolution, we implemented a McDonald-Kreitman (MK) framework based on inStrain-derived polymorphism and substitution statistics^15^. For each gene in each sample, we calculated an MK-style neutrality index as (pN/ps)/(dN/ds), where pN/pS was derived from within-sample polymorphisms (pNpS_variants) and dN/dS from substitutions relative to the reference genome (dNdS_substitutions). Under this formulation, values <1 indicate a relative excess of nonsynonymous substitutions compared with nonsynonymous polymorphisms and were interpreted as suggestive of positive selection in an MK-style framework. Statistical support was evaluated using Fisher’s exact test applied to a 2 × 2 contingency table of nonsynonymous versus synonymous polymorphisms and nonsynonymous versus synonymous substitutions. Genes were considered to exhibit a sample-level positive-selection signal when the neutrality index was <1 and Fisher’s exact test was significant (P ≤ 0.05). For each gene, we calculated (i) the number of selected events, (ii) the number of samples in which the gene was detected, and (iii) the resulting selection frequency.

To further evaluate whether specific genes showed evidence of selection around putative strain-sharing events, we focused on genes that exhibited a positive-selection signal in the recipient host at the first sampling time point when the focal lineage was detected as shared. These genes were interpreted as candidate loci potentially associated with early post-sharing adaptation and were subjected to functional profiling. Functional interpretation of candidate genes was performed using Kyoto Encyclopedia of Genes and Genomes (KEGG) annotation^85^. All genes encoded by the focal genome were mapped to KEGG pathway maps, and pathway assignments for candidate genes were evaluated in the context of neighboring genes and local pathway completeness^85^ (**Table S3**).

### Sharing of ESKAPE and zoonotic pathogens

We followed a previous established protocol of ESKAPE evolutionary analysis^12^. Specifically, for our strains of interest, we jointly analyzed their ANI network as well as phylogenomics(**Table S4**).

#### Analysis of strain sharing by ANI

To gain insight into the genomic similarity of SGB for each participant across species, we calculated pairwise ANI for each species using FastANI with default settings^79^. A custom pipeline was used to plot ANI networks with ggnetwork v0.5.13 in CRAN R (v4.2.2).

#### Analysis of strain sharing by phylogenomics and sequence typing

MAGs and isolate genomes were sequence-typed using MLST v2.23.0 with default settings, for all species with an available PubMLST scheme as of December 2025. For *K. pneumoniae* and other zoonotic bacterial pathogens identified in the ecosystem, phylogenomic trees were reconstructed using GToTree based on translated single-copy marker genes. Publicly available reference genomes matching the sequence types detected in the ecosystem were included for phylogenetic context. For the pathogen trees shown in **Extended Fig. 11**, amino acid alignments of 172 translated single-copy marker genes were used to reconstruct phylogenetic relationships for *Acinetobacter baumannii*, *Staphylococcus aureus*, *Acinetobacter schindleri*, *Bacteroides fragilis*, *Aliarcobacter cryaerophilus*, and *Acinetobacter rivipollensis*, together with matching public reference genomes. MAGs with >50% missing data were excluded, and marker genes were removed when <20% of the gene length was covered. Gaps were retained in the final alignments. Maximum-likelihood phylogenies were inferred using FastTree v2.1.11 as implemented in GToTree and visualized in R using ggtree v3.2.1^86^.

#### K. pneumoniae epidemiology

Given the unexpectedly dominant role of *K. pneumoniae* revealed by MAG-based profiling, we performed culture-based isolation and whole-genome sequencing to further characterize its ecological distribution, population structure, and putative transmission dynamics across the study ecosystem.

#### Isolation of K. pneumoniae from biological samples

A total of 500 biological samples, including human, animal, and environmental specimens, were screened for *K. pneumoniae* isolation. For fecal and soil samples, approximately 0.2g of material was suspended in sterile phosphate-buffered saline (PBS) and vortexed thoroughly. For water samples, aliquots were concentrated by centrifugation or membrane filtration depending on biomass yield, and the resulting pellet or membrane eluate was resuspended in sterile PBS. For swab-based samples, the transport-buffer eluate was used directly. Processed suspensions were inoculated onto selective and differential media for Enterobacterales isolation, including MacConkey agar and - where appropriate - chromogenic media for *Klebsiella* spp., and incubated aerobically at 37 °C for 18–24h. Colonies with morphology consistent with *Klebsiella* spp. were subcultured for purification. Putative *K. pneumoniae* isolates were identified by biochemical testing and further confirmed by 16S rRNA gene sequencing and genome-based taxonomic assignment. When multiple morphologically similar colonies were present on a plate, representative colonies were selected to maximize recovery while minimizing redundancy. Pure isolates were stored in glycerol stocks at −80 °C until DNA extraction.

#### Whole-genome sequencing and annotation

Genomic DNA was extracted from overnight cultures grown in LB broth using a bacterial genomic DNA extraction kit according to the manufacturer’s instructions. DNA quantity and purity were assessed using Qubit fluorometry and NanoDrop spectrophotometry. Sequencing libraries were prepared using a short-read library preparation kit compatible with the Illumina platform and sequenced with paired-end 150-bp reads. Raw reads were quality-filtered and adapter-trimmed using Trimmomatic with default parameters unless otherwise specified^55^. High-quality reads were assembled *de novo* using SPAdes v3.13.1^87^ in --careful mode. Contigs shorter than 500 bp and low-confidence assemblies were excluded where appropriate. Draft genomes were annotated using Prokka v1.14^88^. Species confirmation, MLST, virulence determinants, resistance genes, and capsular loci were determined using Kleborate v2.4.0^89^.

#### Phylogenetic inference

To place the study isolates within the broader diversity of *K. pneumoniae*, publicly available genomes corresponding to the same STs were retrieved from curated databases. Study isolates and ST-matched reference genomes were aligned against a suitable reference genome selected based on sequence type and assembly quality. Reads were mapped using BWA^90^, and SNPs were identified using SAMtools^69^. To minimize phylogenetic distortion caused by homologous recombination, recombinant regions were identified and removed using Gubbins v3.2.8^91^, and only SNPs located in non-recombinant core-genome regions were retained for downstream analyses. Maximum-likelihood phylogenies were reconstructed from the resulting non-recombinant core-genome alignment using IQ-TREE v2.2, with the best-fit substitution model selected by ModelFinder and branch support assessed using 1,000 ultrafast bootstrap (UFBOOT) replicates. To compare phylogenetic structures derived from isolate genomes and MAG-based reconstructions, trees were visualized as tanglegrams. Phylogenies were interpreted in conjunction with associated metadata, including host type, habitat, sampling time, and source compartment, to facilitate the identification of putative transmission links. All phylogenetic visualizations were generated using ggtree v3.2.1^86^.

### Statistical analyses

Unless otherwise specified, statistical analyses are described in the corresponding Methods sections or figure legends.

#### Microbial community diversity and compositional analysis

To evaluate whether sequencing depth and sampling effort were sufficient to capture community richness across habitats, we first examined patterns of γ-diversity. Microbiome richness at genus level and ARG richness at subtype and gene levels were estimated using both observed and asymptotic estimators implemented in iNEXT v3.0.2^22^. ARG γ-diversity, assessed at both the gene and subtype levels, reached asymptotic saturation, indicating that sampling effort was representative across habitats (median completeness, 94.5%; IQR, 92.5–98.1% per habitat; **Extended Fig. 4B**).

Alpha diversity of microbiome and ARG profiles was then quantified using richness, Shannon diversity, and Simpson diversity, as implemented in the R package vegan v2.6.4^92^. Statistical comparisons among habitats were performed using pairwise Wilcoxon rank-sum tests, with P values adjusted by the Benjamini–Hochberg false discovery rate (BH-FDR) procedure. To assess differences in overall community composition, Bray–Curtis dissimilarities were calculated from relative genus abundances, and PCoA was performed using the cmdscale function in R. Sample clustering along the first two PCoA axes was visualized using 95% confidence ellipses and marginal density distributions. Pairwise Bray–Curtis dissimilarities were further summarized to quantify average compositional divergence among habitats and displayed as a dissimilarity heatmap. The contributions of explanatory variables to community variation were assessed using repeated-measures PERMANOVA implemented with the adonis2 function in vegan. Explanatory variables included occupational exposure (farmer versus non-farmer), distance from animals, developmental stage or sampling time, and sample type. The proportion of explained variance (R^2^) was estimated separately for human nasal, human gut, animal, and environmental microbiomes.

To identify features associated with habitat-specific differences, we applied MaAsLin2 v1.8.0^23^ to taxonomic and functional profiles, using arcsine square-root-transformed relative abundances as response variables in generalized linear models. Habitat type and relevant covariates were included as fixed effects, and model coefficients were interpreted relative to predefined reference groups (for example, animal gut, ostrich gut, or goose gut). Statistical significance was determined after Benjamini–Hochberg correction (q < 0.05). This framework was applied consistently across feature types, with reference groups specified according to the biological question.

#### AVD calculation

To quantify the temporal variability of resistome composition across habitats, we calculated the average variability degree (AVD) index following a modified formulation^93^. Lower AVD values indicate greater stability of the community over time.

For each ARG feature i, the variability was defined as the normalized deviation of its abundance across samples:

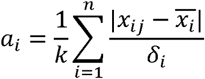

where *x_ij_* represents the abundance of ARG feature *i* in sample *j*, *x̅_i_* is the mean abundance of feature *i* across all samples, δ_*i*_ is the standard deviation, and *k* is the total number of samples.

The overall AVD for each community was then calculated as the average variability across all ARG features:

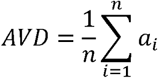

All calculations were implemented in R using a custom script.

#### ARG correlate analyses

Similarly, to quantify differential ARG classes across habitats, we performed multivariable association analyses using MaAsLin2^23^. Arcsine square-root-transformed RPKM-normalized abundances were used as response variables to account for compositionality and heteroscedasticity. Habitat category (for example, farmer gut, non-farmer gut, farmer nasal, non-farmer nasal, and animal hosts) was included as a fixed effect, and individual effect and sampling time was considered as a random effect where longitudinal measurements were available. Model coefficients represent differences in transformed abundance relative to the reference category (non-farming residents). For visualization, mean model coefficients and corresponding standard errors were summarized for each ARG class.

The structural congruence between microbiome and resistome profiles was quantified using Procrustes analysis based on NMDS ordinations of Bray-Curtis dissimilarities derived from genus-level microbial and ARG-subtype abundance matrices. The alignment between the two ordination spaces was evaluated by Procrustes rotation with a PROTEST test implemented in the vegan package. The resulting statistic (m^2^ = 0.3791, p = 0.001) indicated significant global concordance. Each line in the Procrustes plot connects the microbiome and resistome positions of the same sample, where shorter distances reflect stronger structural coupling. Mean Procrustes residuals were computed for each habitat to quantify resistome-microbiome congruence using a custom R script.

A bipartite network was subsequently constructed to reveal habitat-specific enrichment patterns of ARG subtypes. ARG presence was defined as RPKM > 10. For each subtype, Fisher’s exact test was applied to compare occurrence in a focal habitat versus all others, and the Haldane-Anscombe correction was used to estimate the log odds ratio (log OR). Associations meeting BH-FDR < 0.05 and |log OR| > 0.5 were retained for visualization. Edge color and style represented the direction and significance of association (solid red = enriched; dashed blue/green = depleted), and edge width was proportional to |log OR|. Triangles denote habitats and circles indicate ARG subtypes; convex hulls outline ARG neighborhoods around each habitat. Networks were generated using igraph^94^.

#### Permutation-based enrichment of lineage sharing between habitats

To assess whether lineage sharing was preferentially enriched between specific gut-associated and environmental habitats, we implemented a permutation-based framework at the sample-pair level. In each test, the gut habitat was fixed, and only the labels of the competing environmental habitats were permuted. Specifically, this analysis was applied to predefined resident versus non-resident contrasts, including goose gut versus goose soil versus ostrich soil, goose gut versus goose paddling pool versus surrounding river, and ostrich gut versus ostrich soil versus goose gut. Strain-sharing events were first summarized for each unordered sample pair by counting the number of shared genomes identified by inStrain, yielding a sample-pair–level sharing intensity:

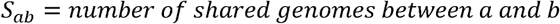

For a focal habitat pair (*i,j*), the observed sharing intensity was defined as the sum of shared genomes across all sample pairs assigned to that habitat combination:

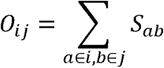

To generate null expectations, we performed permutation tests by randomly shuffling the relevant environmental labels while preserving the underlying sample-pair structure. For each focal comparison, permutations were restricted to a predefined habitat pool consisting of the resident and alternative environmental habitats under consideration. Two permutation schemes were applied: (i) a global model, in which eligible habitat labels were shuffled across all sample pairs, and (ii) a time-stratified model, in which label shuffling was restricted within each sampling time point to control for temporal structure. For each permutation, the reassigned labels were used to recompute *O_ij_*^(*perm*)^, thereby generating a null distribution of sharing intensities under random mixing.

This procedure was repeated 5,000 times. Enrichment was quantified using a z-score:

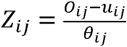

where *u_ij_* and θ_*ij*_ denote the mean and standard deviation of the permuted distribution. One-sided permutation P values were calculated as:

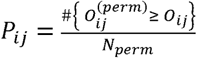

where *N_perm_* = 5,000. Multiple testing correction was performed using the Benjamini–Hochberg procedure within each permutation scheme.

## Notes

### Competing Interest Statement

The authors have declared no competing interest.

### Funding Statement

This study was funded by the Henan Province Key Research and Development Project (Key Project of International Science and Technology Cooperation of Henan Province; grant agreement no. 241111521100) and the Henan Province Science and Technology Research Project (grant agreement no. 252102111013).

