## Extended Fig 1-10 for "Longitudinal cross-species transmission of microbiomes and resistomes across farmers, animals and environment"

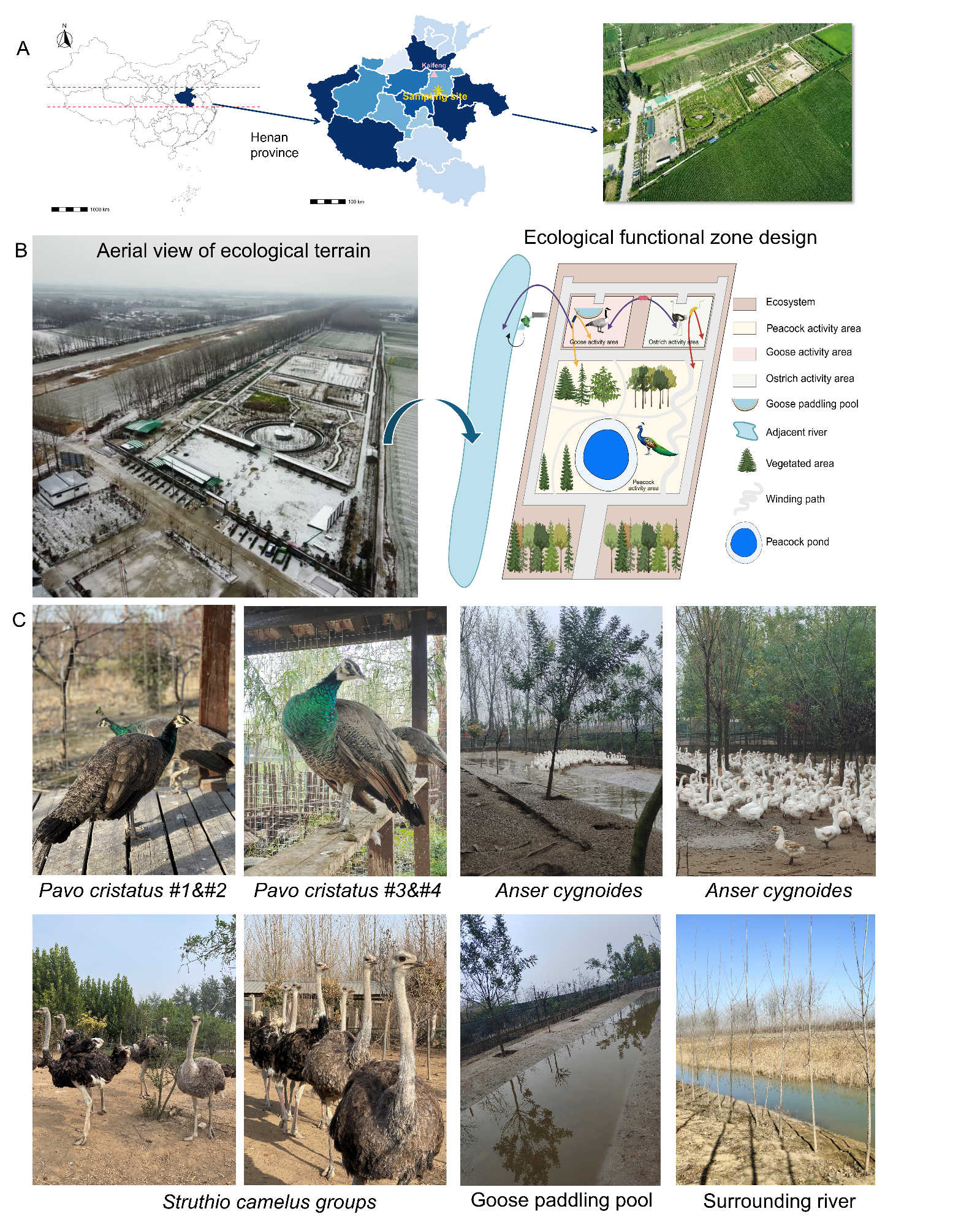

**Extended Fig. S1. Study site, ecological layout and sampling design. A.** Geographic overview of the study site. A three-level map shows the location of the sampling site within Henan Province, China, followed by a zoom-in to the regional scale and an aerial view of the farm ecosystem. **B.** Structural overview of the study system. An aerial photograph of the ecological terrain is shown alongside a schematic diagram of the functional zoning, including host-specific activity areas and environmental compartments (e.g., paddling pool, soil and adjacent river), highlighting potential interfaces for cross-habitat microbial exchange. **C.** Representative images of sampled hosts, environments and field procedures. Photos show animal hosts (*Pavo cristatus*, *Anser cygnoides* and *Struthio camelus*) and environmental habitats (paddling pool and surrounding river).

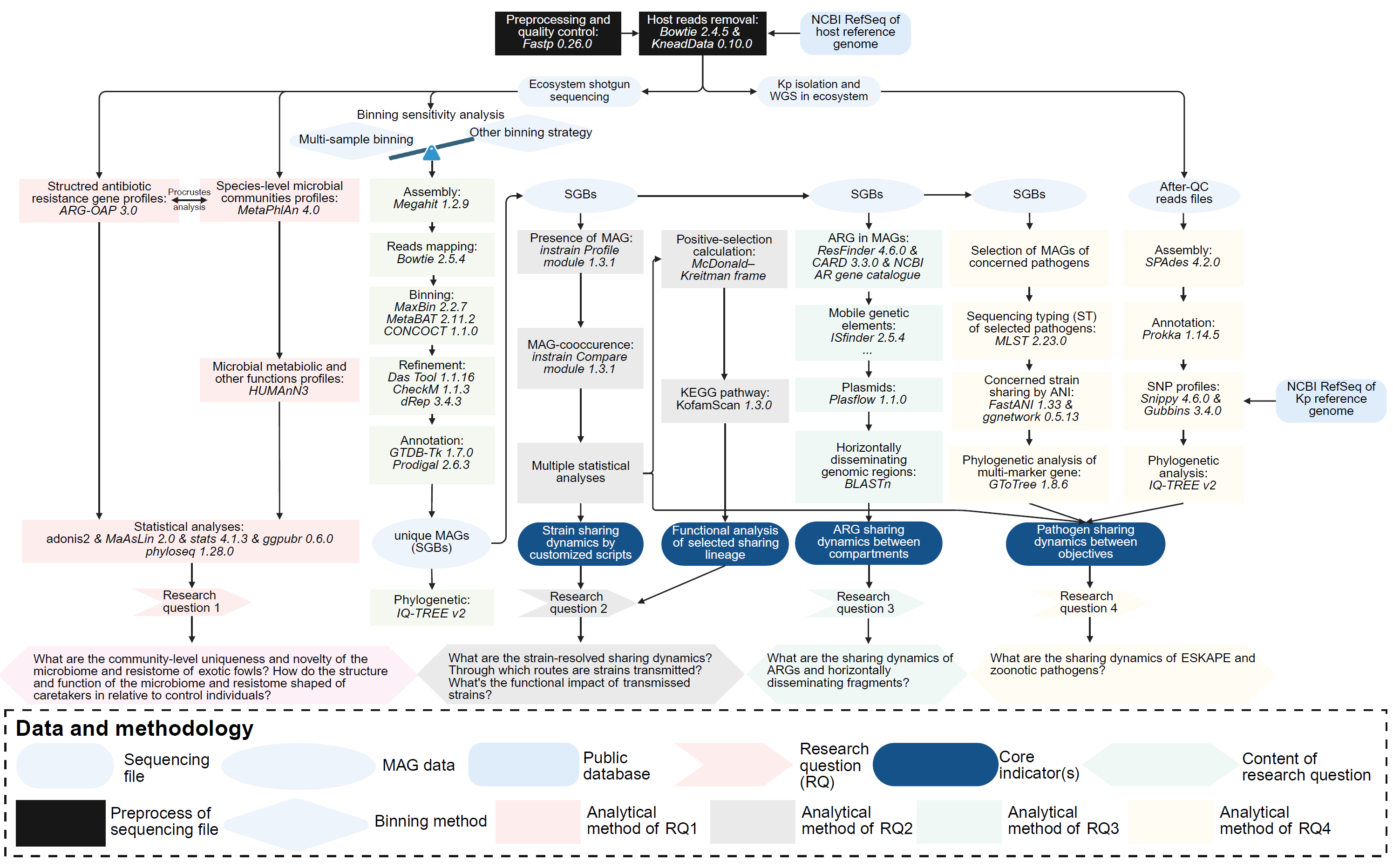

**Extended Fig. S2. Analytic framework.** Diagram summarizing the computational and statistical pipeline, from raw sequencing data processing to metagenomic assembly, binning, genome annotation, taxonomic and functional profiling, strain-sharing analysis, ARG and mobile genetic element analyses, and phylogenetic investigation of selected pathogens.

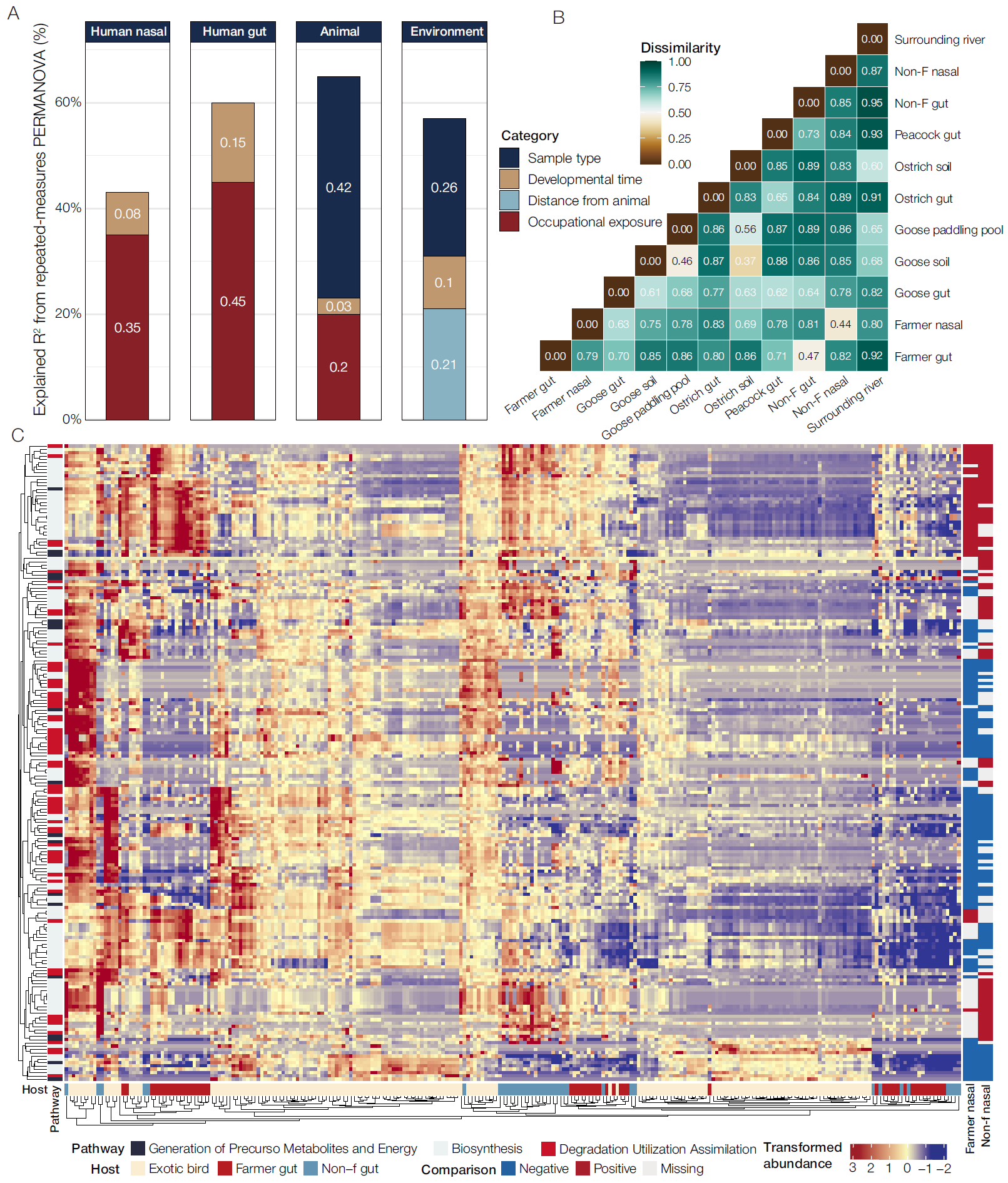

**Extended Fig. S3. Microbial dynamics and metabolism response. A.** Contribution of explanatory factors to microbial community variance across sample types, as assessed by repeated-measures PERMANOVA. Bars indicate the proportion of explained variance (*R^2^*) attributed to occupational exposure, distance from animals, developmental time, and sample type in human nasal, human gut, exotic bird, and environmental microbiomes. **B.** Pairwise Bray–Curtis dissimilarities illustrating average compositional differences among habitats. **C.** Heatmap showing microbial metabolic pathways that differ significantly in relative abundance across host groups, identified using MaAsLin2 (adjusted P < 0.05). Rows represent metabolic pathways and columns represent samples. Color intensity indicates transformed relative abundance from low (blue) to high (red). The left-hand annotation bar indicates pathway functional categories (generation of precursor metabolites and energy, biosynthesis, and degradation/utilization/assimilation). The right-hand annotation bar indicates the direction of significant differential enrichment. Red stars highlight pathways showing evidence of host mixing across groups.

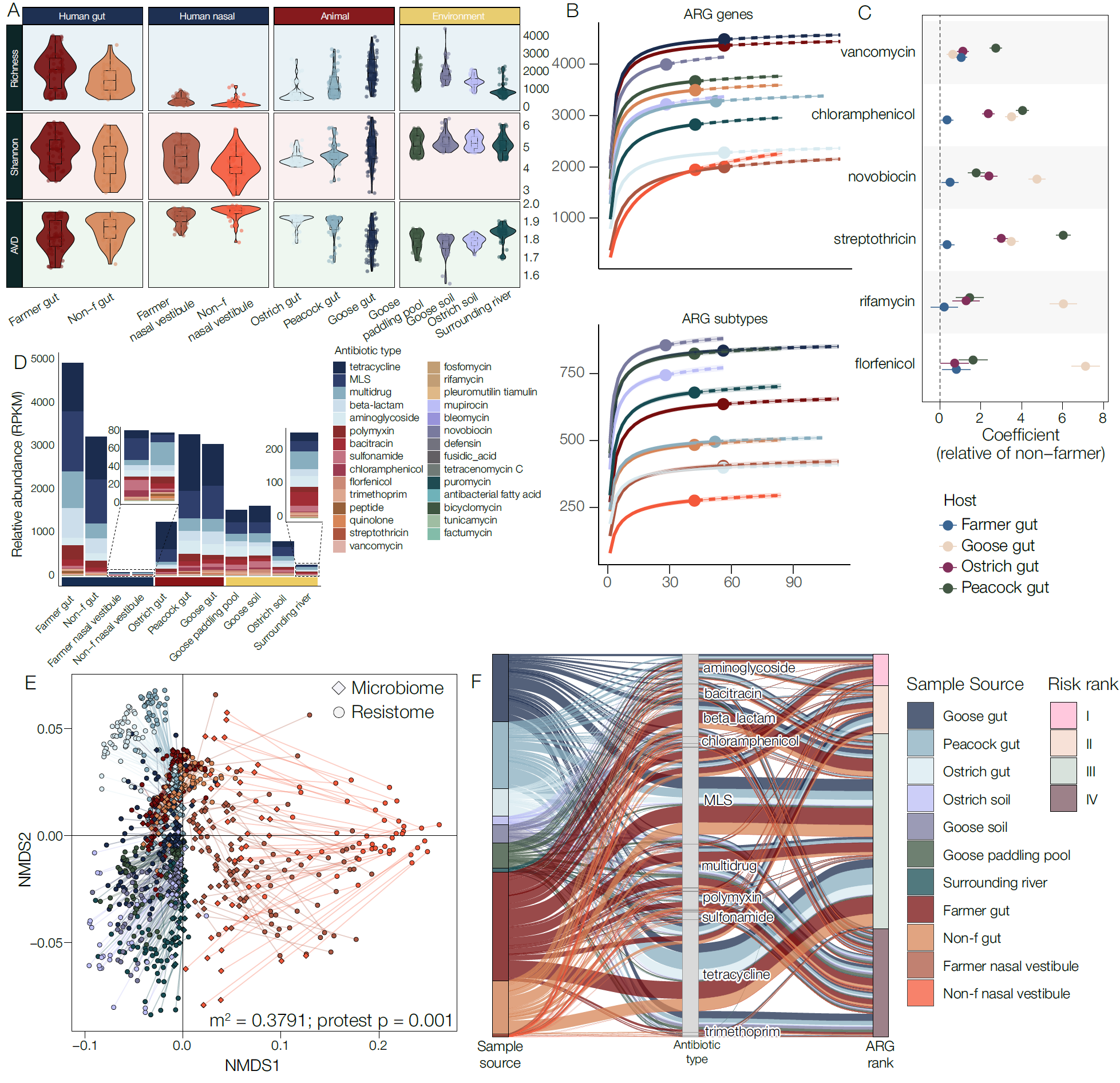
 **Extended Fig. S4. Resistome diversity and compositional structuring across hosts and environments. A.** Alpha diversity of the resistome across 11 sample types, quantified by richness, Shannon diversity and average variation of degree. **B.** Rarefaction curves for ARG richness at the gene and subtype levels across habitats. **C.** MaAsLin2 coefficients for major antibiotic resistance classes in animal gut samples relative to non-farmer gut samples. Points show coefficients and whiskers show standard errors; significant associations were retained after Benjamini–Hochberg correction. **D.** Mean resistome composition across habitats, shown as stacked relative abundances (RPKM) of ARG classes. **E.** Procrustes analysis of microbiome and resistome ordinations across habitats, with paired points representing the same sample (*m*² = 0.3791, PROTEST *P* = 0.001). **F.** Sankey summary of ARG classes and corresponding risk ranks across sample sources.

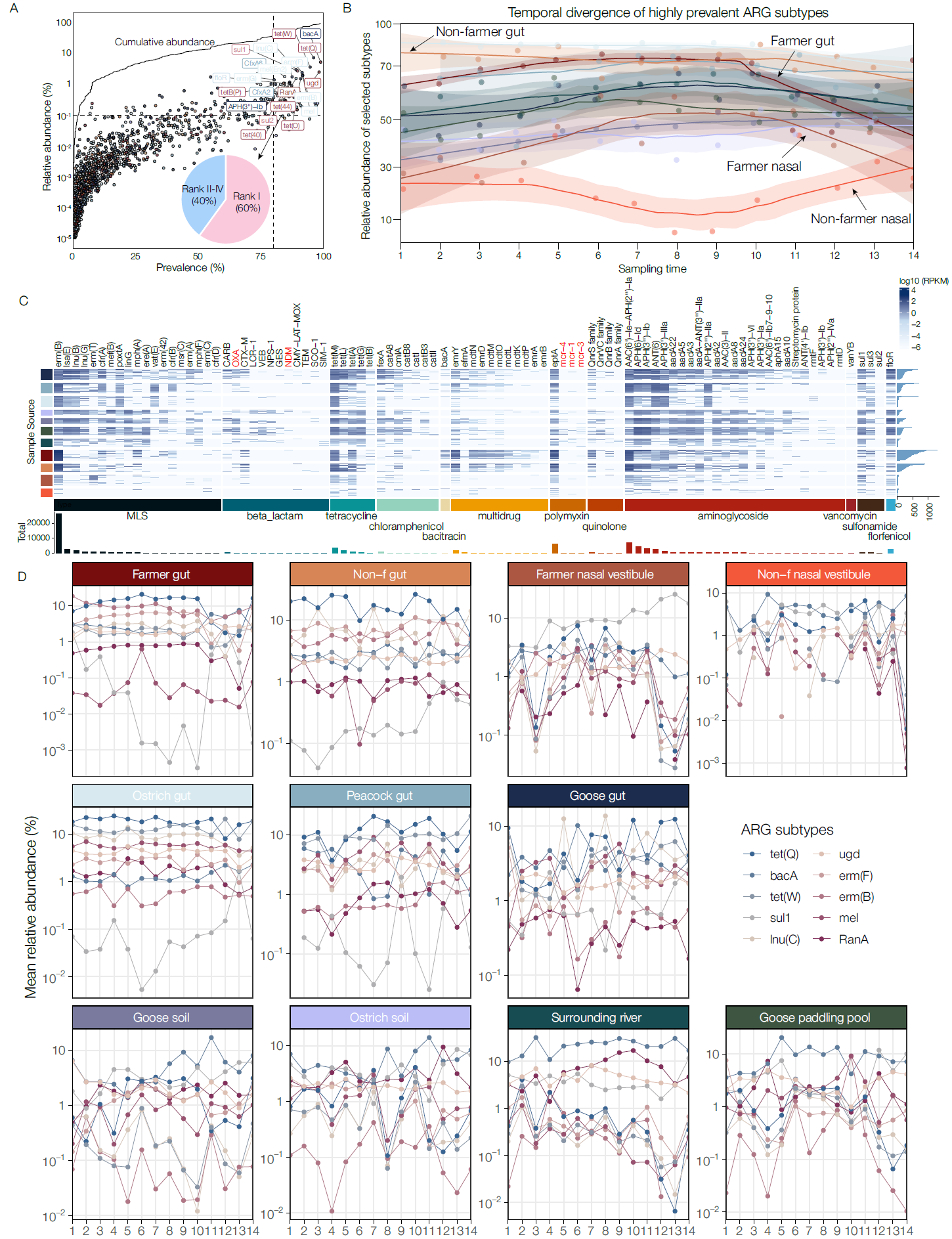

**Extended Fig. S5. Resistome profiling reveals heterogeneous yet interconnected ARG distributions across hosts and environments. A.** Prevalence–abundance landscape of ARG subtypes. Each point represents one ARG subtype, coloured by antibiotic class. The x axis indicates prevalence (percentage of samples with non-zero abundance), and the y axis indicates mean relative abundance (%, log scale). The black curve shows cumulative abundance after ranking ARG subtypes by prevalence. Dashed lines indicate heuristic thresholds for high-prevalence and high-abundance ARGs. Labelled points denote dominant ARG subtypes that jointly exhibited high prevalence and high relative abundance. **B.** Temporal divergence of highly prevalent ARG subtypes across human-associated habitats. LOESS-smoothed curves show temporal changes in the combined relative abundance of ARG subtypes selected using the prevalence–abundance criteria in A. Shaded areas indicate 95% confidence intervals, and points represent mean values at each sampling time. The trajectories show distinct temporal patterns across farmer gut, non-farmer gut, farmer nasal, and non-farmer nasal samples. **C.** Heatmap of the relative abundance of Rank I ARG genes across sample sources. Rows represent sample categories and columns represent ARG genes, grouped by antibiotic class. Colours indicate log-transformed relative abundance (RPKM). The bar plot below summarizes the total abundance of each ARG across the ecosystem. Colours of the left bars correspond to the annotation shown in the header strip of panel D. **D.** Temporal dynamics of the ten most abundant ARG genes across host and environmental compartments. Each panel shows changes in mean relative abundance (%) across sampling time for one sample category.

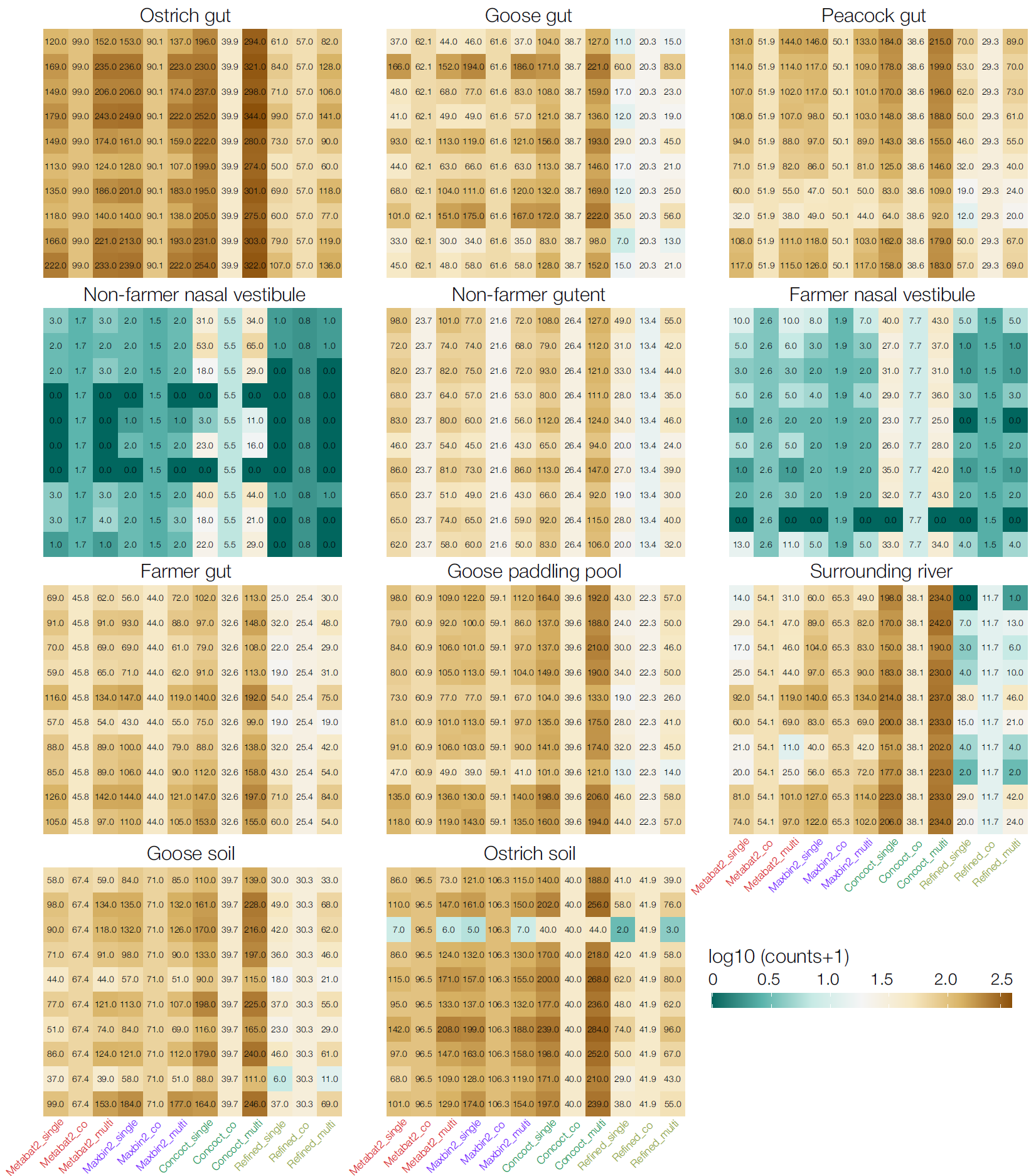

**Extended Fig. S6. Benchmarking of different binning strategies across 11 habitat types.** For each habitat, metagenomic bins were recovered using three representative binning tools under three assembly strategies: single-assembly (_single), co-assembly (_co), and multi-sample binning (_multi). The resulting bins were further refined using MetaWRAP. Bar text colors indicate the corresponding binning tools. For co-assembly, each habitat type yields only one consensus binning result, and the values shown in each cell represent the average across the corresponding batch of samples. In contrast, for single- and multi-sample assemblies, each cell displays the number of bins recovered from the respective individual assembly. Color shading reflects log10 (counts+1). Details of binning modes and tools are described in the Methods.

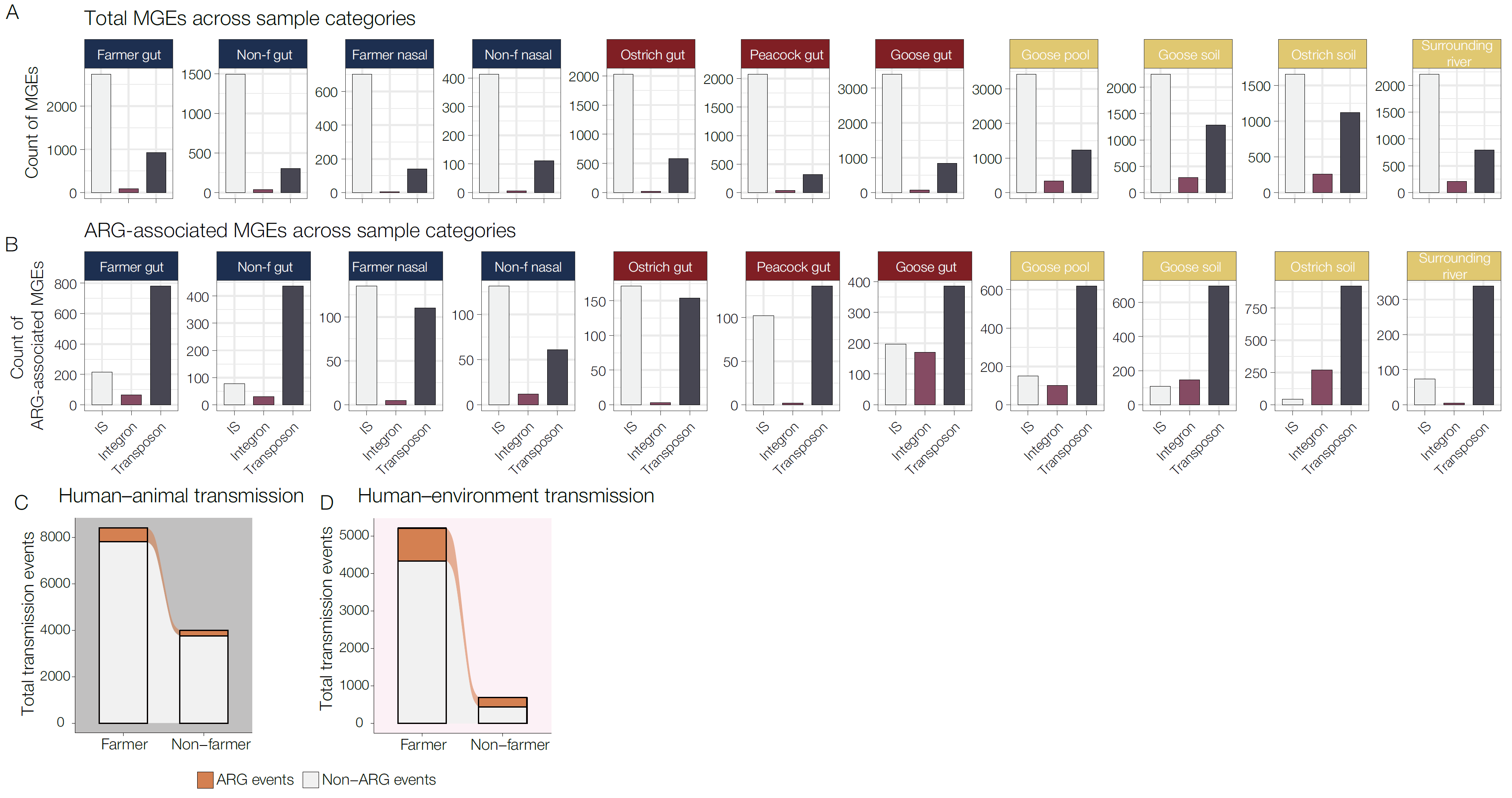

**Extended Fig. S7. Distribution of mobile genetic elements (MGEs) across host and environmental compartments. A.** Total counts of MGEs across sample categories, stratified by MGE class, including insertion sequences (ISs), integrons and transposons. **B.** Counts of ARG-associated MGEs across the same sample categories and MGE classes, highlighting the distribution of resistance-linked mobile elements across host and environmental compartments. **C-D**. Comparison of transmission events between farmers and non-farmers for total HGT events (C) and ARG-related HGT events (D).

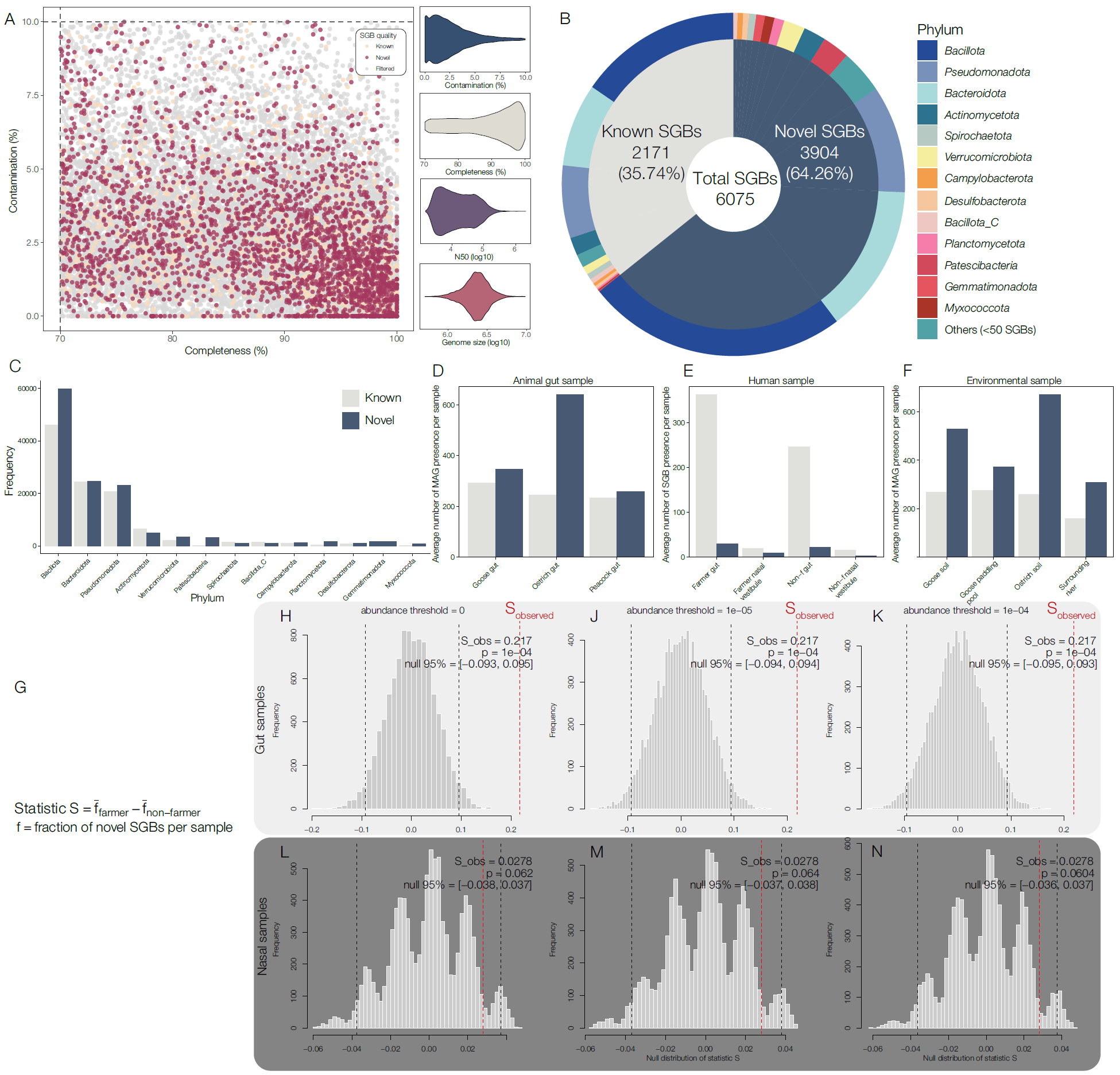

**Extended Fig. S8. Novel SGBs, related to Figure 4 Genome quality, taxonomic composition and distribution of novel species-level genome bins (SGBs) across hosts and environments. A.** Quality profiles of reconstructed metagenome-assembled genomes (MAGs) (n = 29,838), showing completeness and contamination for each genome. Marginal density plots summarize the distributions of contamination, completeness, N50, and genome size. MAGs are coloured by SGB category (known, novel and filtered). **B.** Overall composition of SGBs, showing the proportion of known (n = 2,171) and novel (n = 3,904) SGBs among 6,075 total SGBs, alongside their phylum-level taxonomic distribution. **C.** Phylum-level distribution of SGBs, stratified by known and novel categories, illustrating the contribution of novel diversity within each major lineage. **D-F.** Average number of novel SGBs per sample across host and environmental compartments. (D) Animal gut samples; (E) human-associated samples (farmers and non-farmers); (F) environmental samples, including paddling pool and surrounding river. **G-N.** Permutation-based comparison of the fraction of novel SGBs between farmers and non-farmers. The test statistic was defined as the difference in the fraction of novel SGBs per sample between groups (S = f_farmers − f_non-farmers). Null distributions were generated by random permutation of group labels. Panels show results for gut (H–J) and nasal (L–N) samples under different abundance thresholds. Red dashed lines indicate the observed statistic, and black dashed lines indicate the 95% interval of the null distribution. Results consistently show enrichment of novel SGBs in farmers relative to non-farmers with gut samples showing higher discordance between the groups.

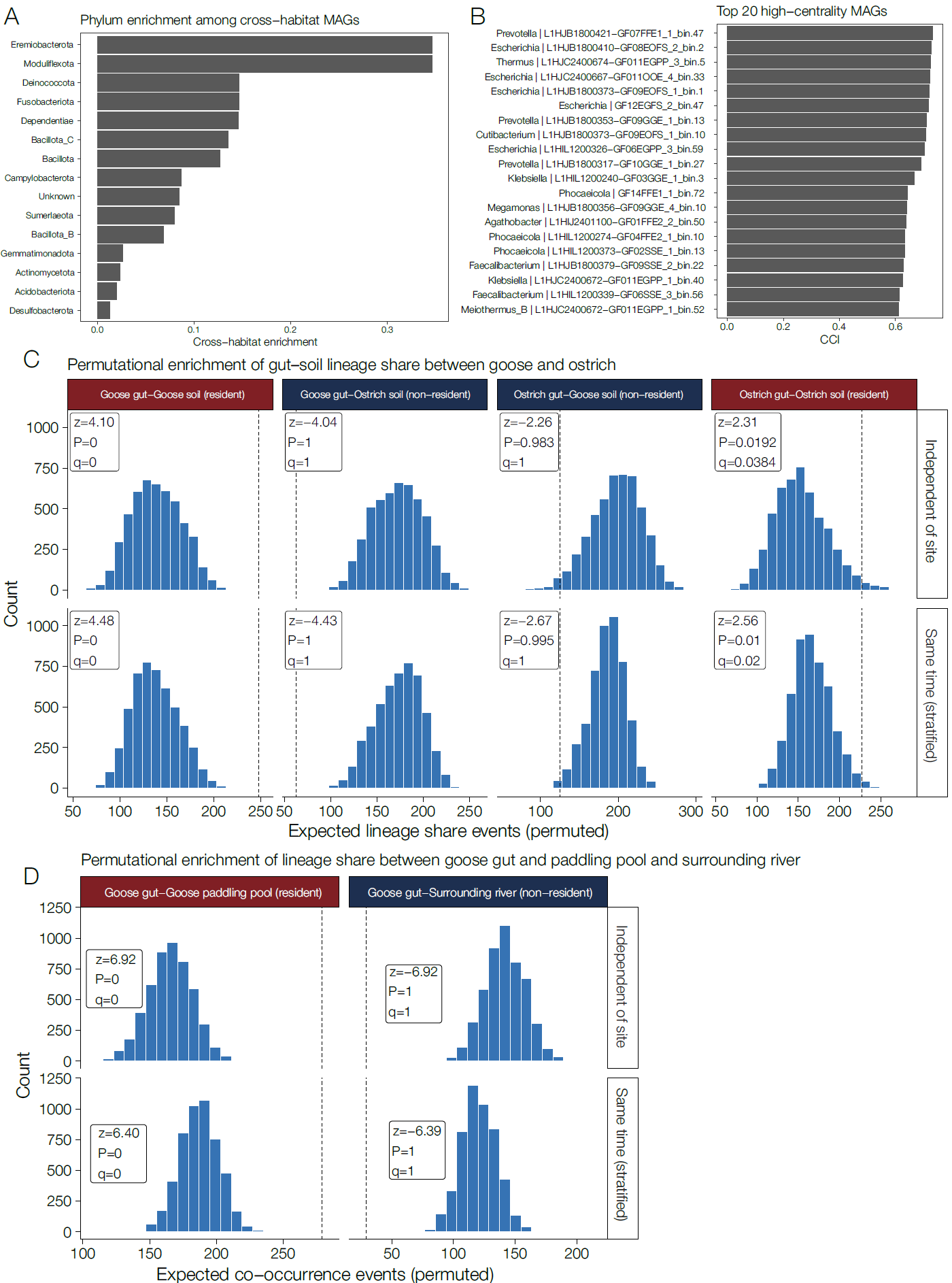

**Extended Fig. S9. Permutation-based enrichment of lineage sharing across gut–environment interfaces. A.** Phylum-level enrichment of MAGs involved in cross-habitat sharing. Bars show the relative enrichment score for each phylum among MAGs detected in strain-sharing events across habitats. **B.** Top 20 MAGs ranked by Cross-habitat Connectivity Index (CCI) in the cross-habitat strain-sharing network. Bars indicate the relative centrality of individual MAGs, highlighting candidate lineages with disproportionate roles in ecological connectivity. **C.** Permutation-based enrichment of lineage sharing between gut and soil habitats in goose- and ostrich-associated compartments. Sharing intensity was quantified as the total number of shared MAGs across sample pairs identified by inStrain. Null distributions were generated from 5,000 permutations under two schemes: a global model, in which habitat labels were shuffled across all eligible sample pairs, and a time-stratified model, in which shuffling was restricted within sampling time points. Resident gut–soil pairs showed significant enrichment of lineage sharing, whereas non-resident combinations did not. **D.** Permutation-based enrichment of lineage sharing between goose gut and aquatic habitats. Sharing was significantly enriched between goose gut and the goose paddling pool, but not between goose gut and the surrounding river, under both global and time-stratified permutation schemes. Together, these results indicate that strain sharing is structured by host-associated habitat coupling rather than random environmental mixing.

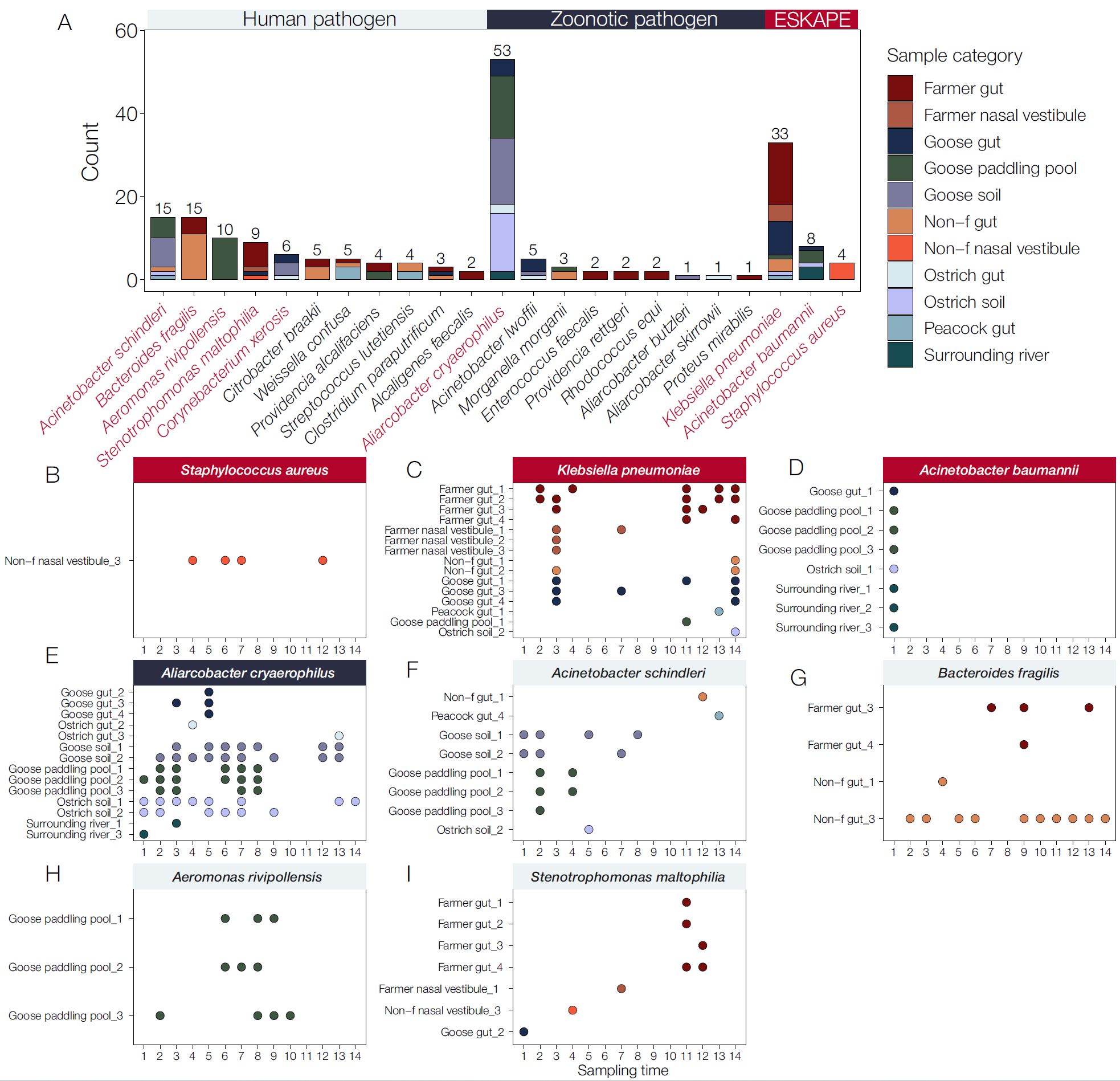

**Extended Fig. S10. MAG recovery of pathogen of concern. A.** Spatiotemporal detection profiles of selected high-priority pathogens, including all ESKAPE species detected in the dataset and the five most prevalent non-ESKAPE pathogens. Each point represents the presence of a given species in a specific sampling ID (defined as sample category × individual ID) at a given sampling time (1–14). Colors indicate sample categories, enabling direct comparison of cross-host and environmental occurrence patterns. **B-I.** Spatiotemporal detection profiles of selected high-priority pathogens, including all ESKAPE species detected in the dataset and the five most prevalent non-ESKAPE pathogens. Each point represents the presence of a given species in a specific sampling ID (defined as sample category × individual ID) at a given sampling time (1–14). Colors indicate sample categories, enabling direct comparison of cross-host and environmental occurrence patterns.

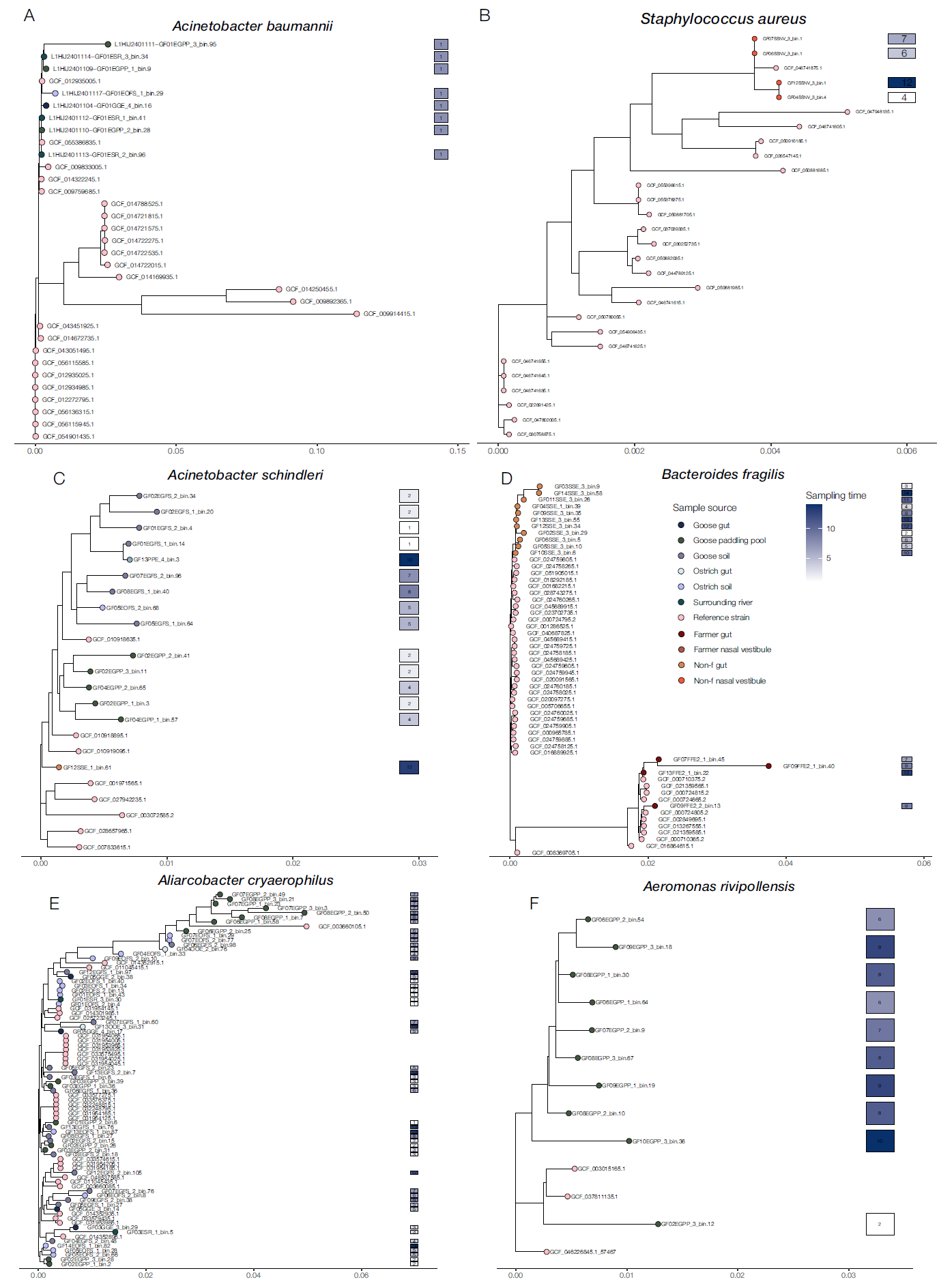

**Extended Fig. S11. Phylogenetic relationships of zoonotic bacterial pathogens identified in the ecosystem.** Phylogenetic trees were reconstructed from amino acid alignments of 172 translated single-copy marker genes for 8 *A. baumannii* MAGs **(A)**, 4 *S. aureus* MAGs **(B)**, 15 *A. schindleri* MAGs **(C)**, 15 *B. fragilis* MAGs **(D)**, 53 *Aliarcobacter cryaerophilus* MAGs **(E)**, and 10 *A. rivipollensis* MAGs **(F)**, together with publicly available reference genomes matching the sequence types identified in the ecosystem. Tip-point colors indicate strain source. The boxes shown to the right of each tip indicate sampling time.
