## Supplementary Note 1-4 and Supplementary Fig 1-3 for "Longitudinal cross-species transmission of microbiomes and resistomes across farmers, animals and environment"

**Supplementary Notes**

***Supplementary Note 1: Basic information on the sampled ecosystem***

The ecosystem housed four peacocks, ten ostriches, and thirty geese during the study period. Peacocks and geese were introduced into the ecosystem in 2023, whereas ostriches were maintained on the farm since 2021. In addition, four resident workers were regularly involved in animal husbandry activities, primarily feeding and caring for all animals. These workers typically returned to their homes for 1–2 days per week and resided on the farm for the remainder of the time, thereby constituting the human population with the most sustained exposure to the animal-associated environment. To maximize representation of environmental heterogeneity, soil samples were collected from multiple directions and depths within animal activity areas at each sampling time point. This design was intended to capture spatial variation in soil-associated microbiomes as comprehensively as possible (see **Methods**).

***Supplementary Note 2: Sensitivity analysis of metagenomic binning approach***

To assess the robustness of MAG recovery across heterogeneous habitats, we benchmarked three binning strategies, i.e., single-sample assembly, co-assembly, and multi-sample binning, across 11 ecological compartments (**Extended Data Fig. S6**). Multi-sample binning consistently outperformed alternative approaches, yielding the highest number of recovered bins across all habitats. This effect was particularly evident in complex animal-associated and environmental microbiomes, where multi-sample binning markedly increased genome recovery relative to both single-sample and co-assembly strategies. In contrast, co-assembly approaches resulted in reduced bin counts, consistent with the merging of closely related strains during joint assembly. Single-sample assembly showed moderate performance but exhibited substantial variability across habitats. Notably, these trends were consistent across different binning tools, suggesting that assembly strategy, rather than tool choice, is the primary determinant of genome recovery efficiency. Together, these results support the superiority of multi-sample binning for reconstructing strain-resolved genomes in complex, multi-host ecosystems, providing a strong methodological foundation for its use in downstream transmission analyses.

***Supplementary Note 3: Mapping the metabolic heterogeneity across habitats***

Given partial convergence in taxonomic composition across habitats, we next assessed whether our observed microbiome relationships were reflected at the functional level. We used HUMAnN3 to characterize metabolic potential in human (farmers and non-farmers) and animal (goose, peafowl and ostrich) shotgun metagenomic samples^1^. Consistent with the compositional patterns, pathway mapping rates differed significantly across habitats. In humans, gut metagenomes showed significantly lower pathway mappability than nasal metagenomes, and farmers consistently exhibited lower annotation rates than non-farmers in both body sites (**Supp. Fig. S2 D-F**). Animal-associated microbiomes showed higher proportions of unmapped reads overall, with ostrich gut microbiomes displaying the highest unmapped fraction and goose gut microbiomes the lowest (**Supp. Fig. S2 A-B**). These results suggest that a substantial fraction of metabolic potential in animal-associated communities, particularly in ostriches, remains poorly represented in current reference databases. Despite these annotation differences, clear ecological structuring was also evident at the level of annotated metabolic pathways. Among gut-associated communities, farmers exhibited significantly higher pathway diversity than non-farmers (**Supp. Fig. S2D-E**), whereas geese showed the highest Shannon diversity among the three animal hosts (**Supp. Fig. S2F**). Bray–Curtis PCoA of annotated pathways showed that farmer gut microbiome overlapped more extensively with animal gut metabolic profiles than did non-farmer gut microbiomes, while annotated pathways from human nasal samples clustered adjacent to the gut-associated β-diversity space, forming a metabolically redundant region (**Supp. Fig. S2C**). These findings suggest that, despite substantial taxonomic divergence across hosts, part of their cumulative metabolic potential converges.

We then used MaAsLin2 to identify differentially enriched pathways across habitats. Although animal guts, farmer samples and non-farmer samples each retained many habitat-specific pathways, we also identified a subset of positively enriched pathways showing significant cross-habitat overlap^2^ (**Fig. 2E**). These shared pathways clustered more closely with farmer gut microbiome and were primarily related to degradation, utilization and assimilation as well as biosynthetic functions, suggesting that metabolic convergence may be driven by ecologically shared taxa exchanged between animals and farmers. This pattern was even more significant in comparisons between farmer nasal microbiome and animal gut communities (**Extended Fig. S3C**), further supporting the unstable and exposure-sensitive nature of the nasal microbiome^3^. Together, these results indicate that long-term occupational exposure promotes convergence of the taxonomic and functional metabolic components of both the gut and nasal microbiome of farmers toward animal-associated communities.

***Supplementary Note 4: Novelty of microbiome in this ecosystem***

Taxonomic profiling based on MetaPhlAn4 first suggested that this ecosystem harbored a high degree of microbial novelty (**Fig. 1A**). Taxonomic analysis of bacterial MAGs further confirmed this pattern, revealing extensive novelty across the reconstructed genome catalog, including many lineages detected at relatively high abundance. Based on the assembled species-level genome bin (SGB) catalog, we found that 3,904 of 6,075 unique SGBs (64.26%) lacked a reference genome within 95% average nucleotide identity (ANI), indicating that they likely represent previously undescribed species. These were hereafter defined as unknown SGBs (uSGBs) (**Fig. 4B; Extended Data Fig. S8B**). These uSGBs were distributed across multiple bacterial phyla, with the largest fractions belonging to Bacillota (including *Bacillota_A* and *Bacillota_C*), Bacteroidota, and Pseudomonadota (**Extended Data Fig. S8 B,C**). Notably, farmer-associated MAG collections showed greater taxonomic novelty than those from non-farming residents (**Extended Data Fig. S8E**). Permutation-based analyses further revealed a strong site-specific pattern in the enrichment of novel MAGs. In nasal samples, farmer-associated microbiomes contained a substantially higher fraction of novel MAGs than non-farmer samples (Δ = 0.217), a difference that was highly significant (P = 1 × 10^-4^) and robust across a range of abundance thresholds. In contrast, gut samples showed a smaller but consistent increase in novel MAG fraction among farmers (Δ ≈ 0.028), although this difference did not reach statistical significance (P ≈ 0.06) regardless of abundance filtering (**Extended Data Fig. S8G–N**). Among animal hosts, ostriches exhibited the greatest burden of unknown MAGs. This pattern was also reflected in the corresponding animal-associated soils, which harbored elevated numbers of uSGBs, suggesting that animal occupancy may contribute to reshaping environmental microbial composition (**Extended Data Fig. S8D–F**). By comparison, the surrounding river, used here as an external environmental control, contained the lowest proportion of uSGBs. Together, these observations raise the possibility that sustained animal occupancy substantially alters both host-associated and surrounding environmental microbiomes within the ecosystem. Consistent with these findings, maximum-likelihood phylogenies of representative SGBs showed that uSGBs were broadly distributed across the bacterial tree and substantially expanded known phylogenetic diversity (**Fig. 4B**). This pattern further underscores how underexplored host-associated ecosystems can provide an important window into previously unrecognized microbial diversity and host–microbiome co-evolution.

**Supplementary Figures**

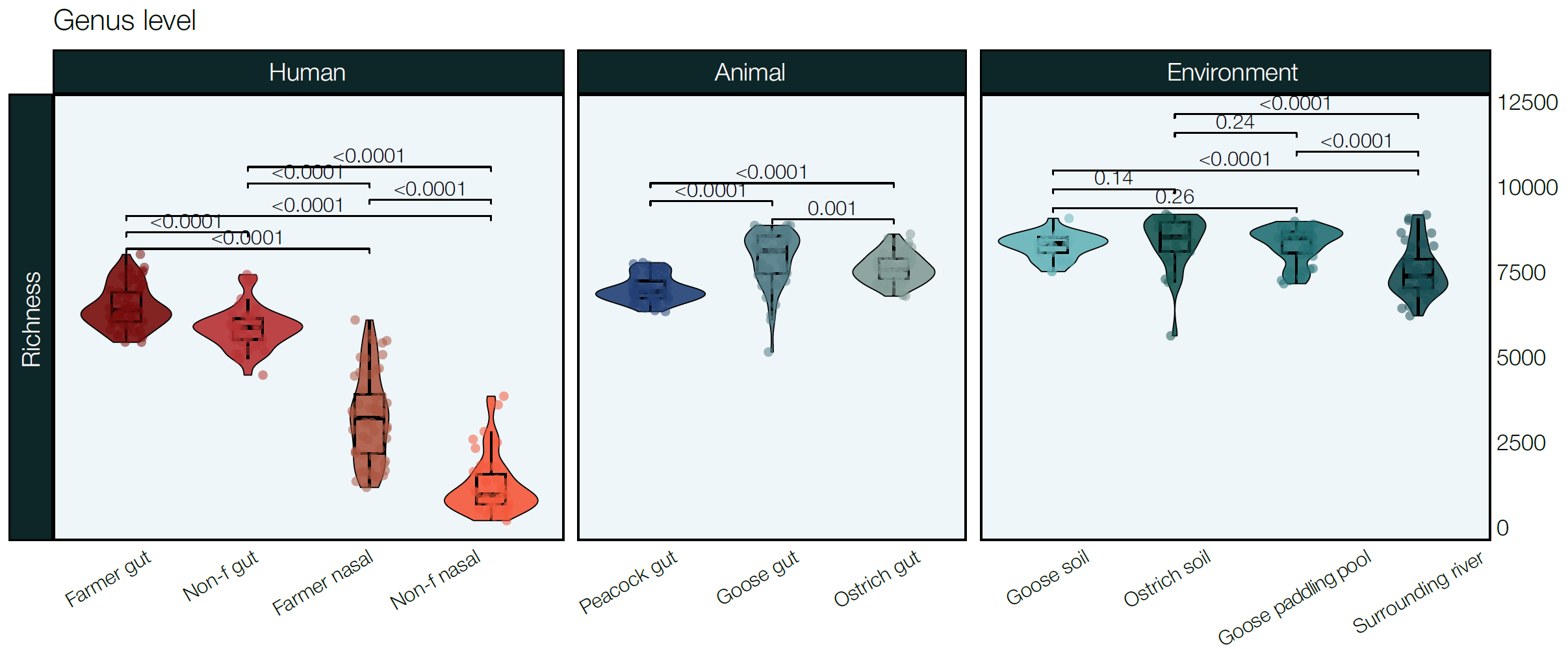

**Figure S1. Violin plots showing genus-level richness across habitats.** Samples are grouped by major habitat categories (human, animals, and environment) and further stratified by specific sampling sites. Horizontal lines indicate median values.

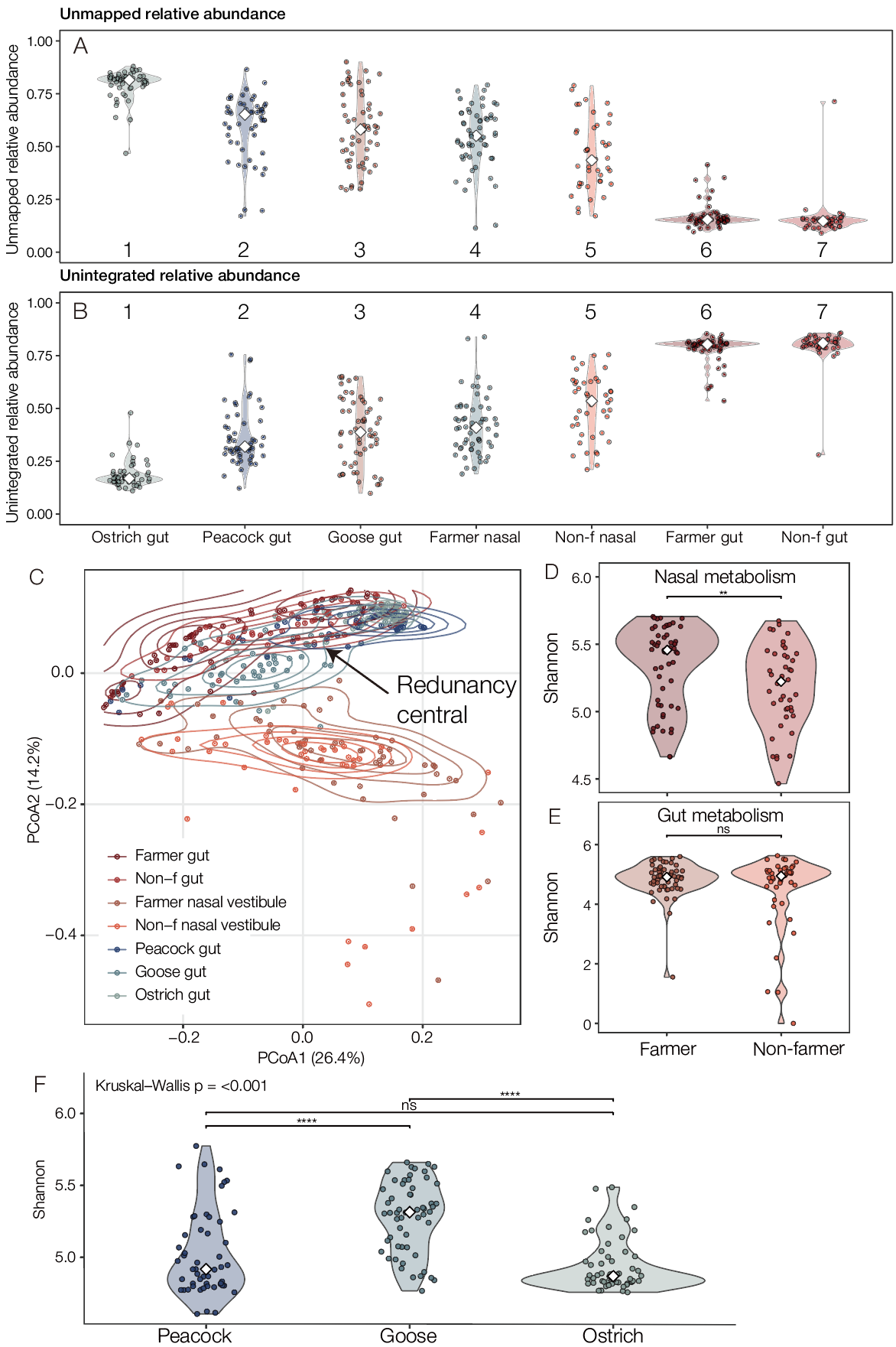

**Figure S2. A.** UNMAPPED relative abundance represents reads that cannot be aligned to any known gene family and therefore reflect the extent of reference database coverage. **B.** UNINTEGRATED relative abundance represents gene families that were successfully annotated but not incorporated into any pathway during reconstruction, reflecting the stringency and specificity of HUMAnN3’s pathway integration process. **C**. PCoA based on Bray-Curtis dissimilarity of inferred microbial metabolic pathway profiles across host groups. **D-F**. Violin plots showing Shannon diversity of microbial metabolic pathways (inferred using HUMAnN3) across nasal samples (**D**), gut samples (**E**), and animal gut samples (**F**). Statistical comparisons were performed using the Kruskal-Wallis test.

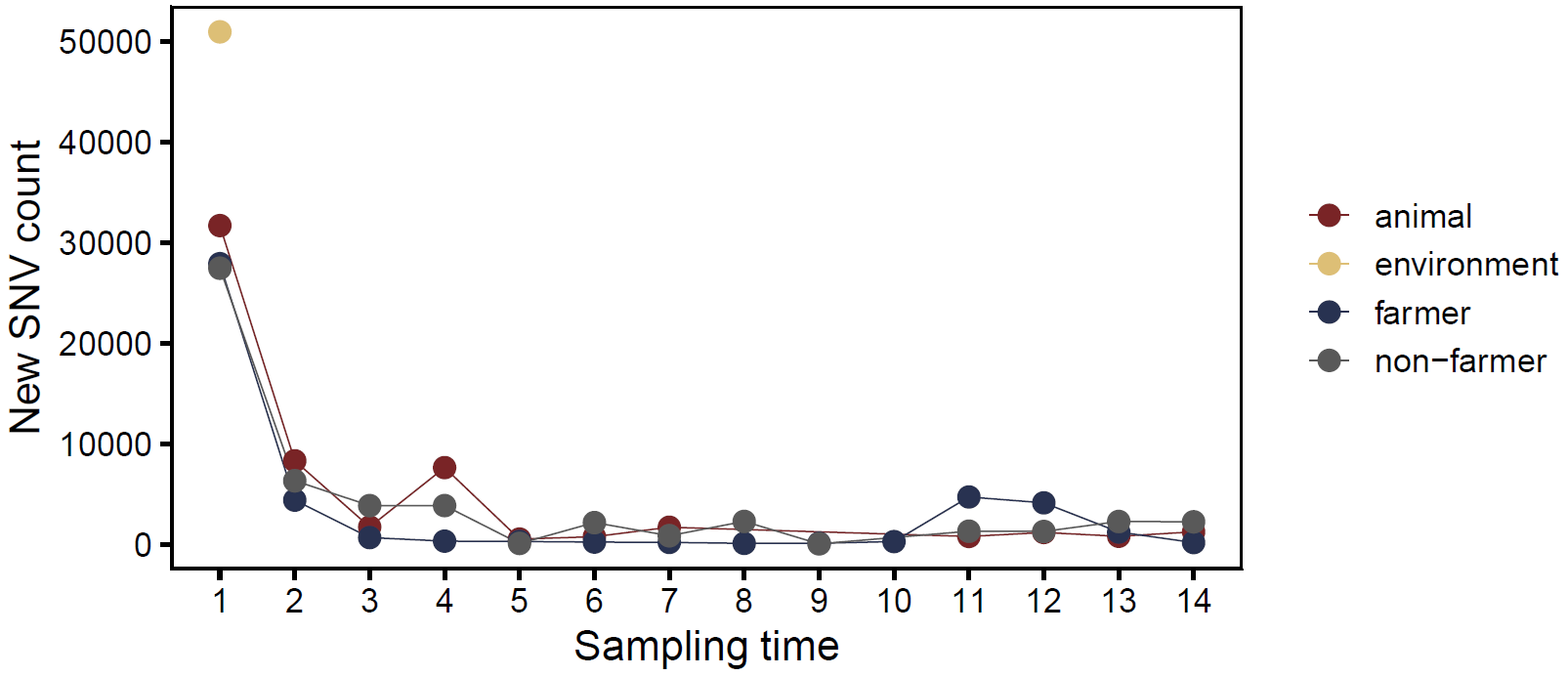

**Figure S3. The introduction of new SNV counts of *E. coli GF12EGFS_2_bin.26*.** The absence of a spike in the number of new SNVs suggests that an increase in the evenness of existing strains occurred, rather than the introduction of new strains.
